# Genome-wide analyses of quantitative generalised anxiety symptom severity

**DOI:** 10.1101/2025.07.10.25331321

**Authors:** Megan Skelton, Brittany L Mitchell, Elham Assary, Danyang Li, Genevieve Morneau-Vaillancourt, Alan E Murphy, Abigail R ter Kuile, Rujia Wang, Mark J Adams, Enda M Byrne, Elizabeth C Corfield, Poppy Z Grimes, Laurie J Hannigan, Jihua Hu, Kadri Kõiv, Alex SF Kwong, Sergi Papiol, Johanne H Pettersen, Giorgio Pistis, Enrique Castelao, Nora I Strom, Peter J van der Most, Anxiety Disorders Working Group of the Psychiatric Genomics Consortium, GLAD+ authors, Lifelines Cohort Study, NIHR BioResource, Protect-AD Consortium, Ole A Andreassen, Angelika Erhardt-Lehmann, Alexandra Havdahl, Nathan Skene, Brad Verhulst, Heike Weber, Chérie Armour, Helga Ask, William E Copeland, Udo Dannlowski, Jürgen Deckert, Katharina Domschke, Ian B Hickie, Kelli Lehto, Tina B Lonsdorf, Ulrike Lueken, Michelle K Lupton, Sarah E Medland, Andrew M. McIntosh, Albertine J Oldehinkel, Martin Preisig, Andreas Reif, Harold Snieder, James T. R. Walters, Naomi R Wray, Catharina A Hartman, Nicholas G Martin, John M Hettema, Gerome Breen, Jonathan RI Coleman, Thalia C Eley

**Author notes:** Joint first authors contributed equally. Senior authors contributed equally. Corresponding author: Prof Thalia C Eley.

## Abstract

We performed a genome-wide association meta-analysis of generalised anxiety symptom severity in 693,869 individuals of European ancestry from 14 cohorts. We identified 80 independent genome-wide significant variants within 74 loci, 39 of which were novel for anxiety. SNP-based heritability was 5.9% (SE = 0.15%). Polygenic scores were significantly associated with anxiety symptom severity and disorder in European, African, and South Asian ancestry samples (r^2^=1.2%-2.9%). Significant genetic correlations were estimated with numerous mental and physical health traits, including case-control anxiety, neuroticism and depression (r_g_=0.71-0.86), irritable bowel syndrome (r_g_=0.57), coronary artery disease, endometriosis, and migraine (r_g_=0.20-0.27). Gene-based and pathway analyses implicated synaptic and axonal processes, with enriched expression in the brain. These findings highlight the additional value of a quantitative approach in anxiety genetics.

## Introduction

Anxiety disorders are the most prevalent mental health conditions worldwide^1^ and rates are rising^2–4^. Anxiety is associated with reduced quality of life^5^, elevated mortality^6,7^, and is frequently comorbid with other mental^8–12^ and physical^13^ health conditions, from irritable bowel syndrome to cancer. When co-occurring with other conditions, anxiety symptoms can exert an independent and sometimes greater impact on quality of life and functioning than the primary diagnosis^8^, as has been reported in autism^14^ and bipolar disorder^15^.

Twin and family studies estimate the heritability of anxiety disorders at 20-60%^13^, with measures capturing stable anxiety typically showing higher heritability^16^. Early case-control genome-wide association studies (GWAS), which aggregated across anxiety subtypes, identified a handful of risk loci^17–19^. More recent large-scale efforts employing a range of analytical approaches have reported between 14 and 51 associated loci^20–22^. The largest GWAS of anxiety cases to date, also by the Anxiety Disorders Working Group of the Psychiatric Genomics Consortium (PGC-ANX), identified 58 independent loci from over 120,000 cases^23^. Across these studies, single nucleotide polymorphism (SNP)-based heritability estimates ranged from 5% to 10%^20,22,23^.

Fear and worry serve an evolutionary function by promoting vigilance and caution in response to potential threats^24^. Variation in threat sensitivity across individuals may be adaptive at the group level, and consistent with this, anxiety symptoms exist in the population along a continuum of frequency and severity. Clinical anxiety represents a practical threshold at the upper extreme of this distribution, based on levels of distress and functional impairment. GWAS of anxiety using quantitative symptom scores capture genetic variation across the full phenotypic range, not only at clinical thresholds. This approach can offer greater statistical power^25^ and a more comprehensive representation of genetic influences on anxiety traits. The degree of genetic overlap between quantitative anxiety symptom severity and disorder status has received little study. A GWAS of a two-item anxiety scale in the European-ancestry subsample of the Million Veteran Program (MVP) identified five significant loci^26^, and moderate-to-strong genetic correlations with lifetime anxiety disorder (r_g_ = 0.59 - 0.87^13,26^). However, the brevity of the measure may have limited the potential to comprehensively capture phenotypic variance. Additional support for shared genetic influences across the anxiety continuum, including above and below diagnostically relevant thresholds, comes from a UK Biobank study reporting high genetic correlations between mild, moderate, and severe anxiety symptom groupings (r_g_ = 0.76 - 0.98^19^). Among individuals with lifetime anxiety or depression, anxiety symptom severity has also shown a significant genetic correlation with functional impairment ratings (r_g_ = 0.79^27^), which have clinical relevance.

These findings provide preliminary evidence that a well-powered GWAS of quantitative anxiety would identify variants relevant to both symptom severity and clinically defined anxiety. Besides the MVP analysis of a two-item scale, previous quantitative anxiety GWAS have incorporated other psychiatric or personality traits to maximise statistical power^28,29^, resulting in findings relating to a broader construct than anxiety symptoms specifically. To address this, we performed the largest genome-wide association meta-analysis of generalised anxiety disorder (GAD) symptom severity to date. We analysed data from 693,869 individuals of inferred European ancestry across 14 cohorts from PGC-ANX. Most cohorts used the Generalised Anxiety Disorder 7-item scale (GAD-7) or conceptually similar self-report measures of recent GAD symptoms. In addition to identifying associated loci, we performed variant- and gene-level investigations, estimated genetic correlations with relevant traits, and evaluated polygenic score prediction in independent samples of European, African, and South Asian ancestry.

## Results

### Genome-wide association meta-analysis

We performed a genome-wide association meta-analysis of GAD symptom severity in 693,869 individuals of European ancestry across 14 cohorts from eight countries (see Tables S1-S3 for cohort, phenotype, and GWAS details). The analysis identified 80 independent genome-wide significant variants (*p* < 5×0^−8^) across 74 loci (Fig 1, Fig S1, Table S4). The top signal came from an intergenic locus near the long noncoding RNA *RP4-598G3*.*1* on chromosome 1 (lead SNP rs7546305, *p* = 3.8 × 10^−15^, beta = −0.014, SE = 0.0017), followed by a locus within an intron of *PCLO* on chromosome 7 (lead SNP rs1476548, *p* = 4.9 × 10^−15^, beta = −0.014, SE = 0.0018). These loci have both previously been associated with internalising traits, including anxiety (Table S5).

**Fig. 1.**
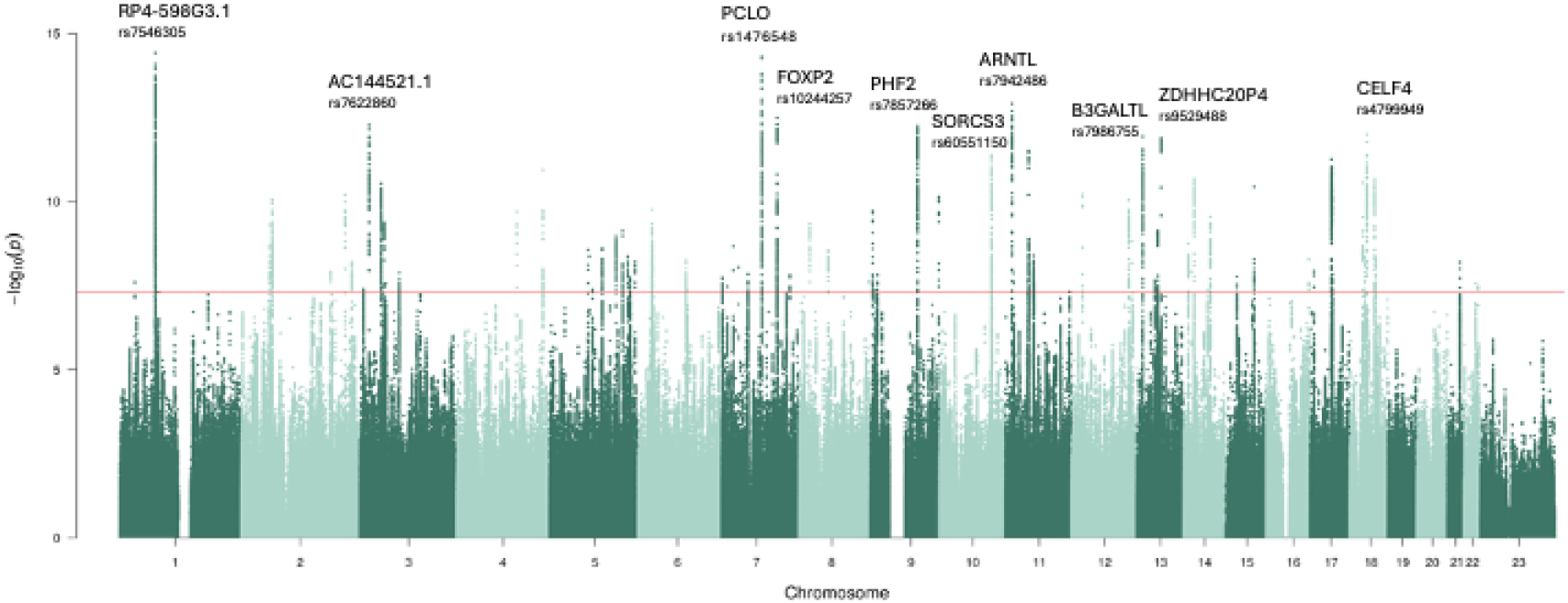
Manhattan plot of the genome-wide association meta-analysis of generalised anxiety symptom severity (N = 693,869). Variants are represented as points, plotted against their respective genomic location and association significance value. The red line indicates the genome-wide significant threshold (*p* < 5×0^−8^).

Of the 74 loci, 16 had no prior associations with internalising trait GWAS, including neuroticism, anxiety, or depression, at any variant in linkage disequilibrium (LD) with the lead SNP (r^2^ > 0.1; Methods, Table S5a). Thirty-nine loci were novel for anxiety specifically, measured either as a diagnosis or symptom severity phenotype. Of the 58 loci previously identified in the PGC-ANX anxiety disorder study^23^, all showed the same direction of effect in the present analysis, with 19 (33%) reaching genome-wide significance, and a further 33 (57%) showing nominal significance (*p* < 0.05) (Table S5b). Nine of the 14 cohorts included here also contributed to the PGC-ANX anxiety disorders study, although cohort sample composition somewhat differed due to the availability of symptom score versus diagnostic information. Power calculations^30^ confirmed increased power in the present analysis relative to the anxiety disorder study (Table S6).

Among the 6,012 genome-wide significant SNPs, study heterogeneity accounted for a moderate proportion of variance in effect size estimates, with a median I^2^ across cohorts of 29.7%. In contrast, the median I^2^ for the 80 independent genome-wide significant SNPs was 0%, reflecting highly consistent effects across cohorts. Heterogeneity statistics for the lead SNPs are presented in Table S4. Genetic correlations between sufficiently powered cohorts, estimated using linkage disequilibrium score regression (LDSC), ranged from 0.64 to 0.97 (Table S7). As a sensitivity analysis, we categorised cohorts by generalised anxiety symptom severity measure (e.g. GAD-7; six subgroups) and by ascertainment method (‘community’ and ‘clinical’ subgroups) (Table S2). Subgroup meta-analyses were performed as per the main analysis, and genetic correlations between subgroups were estimated with LDSC. While most subgroup comparisons were underpowered, including that for the ascertainment method, the genetic correlation between the two most frequently used measures, GAD-7 and GAD-2, was high (r_g_ = 0.86, SE = 0.031; Table S8).

### SNP-based heritability and genetic correlations with external traits

The SNP-based heritability estimate from SBayesRC was 5.90% (SE = 0.146%). SBayesRC was selected due to the known under-estimation of heritability from LDSC^31^. LDSC^32^ indicated that genomic inflation was largely not attributable to confounding (intercept = 1.03, SE = 0.010). To characterise the broader genetic architecture of GAD symptoms, we used LDSC^33^ to estimate genetic correlations with 105 traits spanning mental and physical health, personality, cognitive, and socioeconomic domains (Table S9). Following Bonferroni correction (*p* < 4.76 ×0^−4^), 64 associations remained significant (Fig 2, omitting four traits represented by multiple studies).

**Fig. 2.**
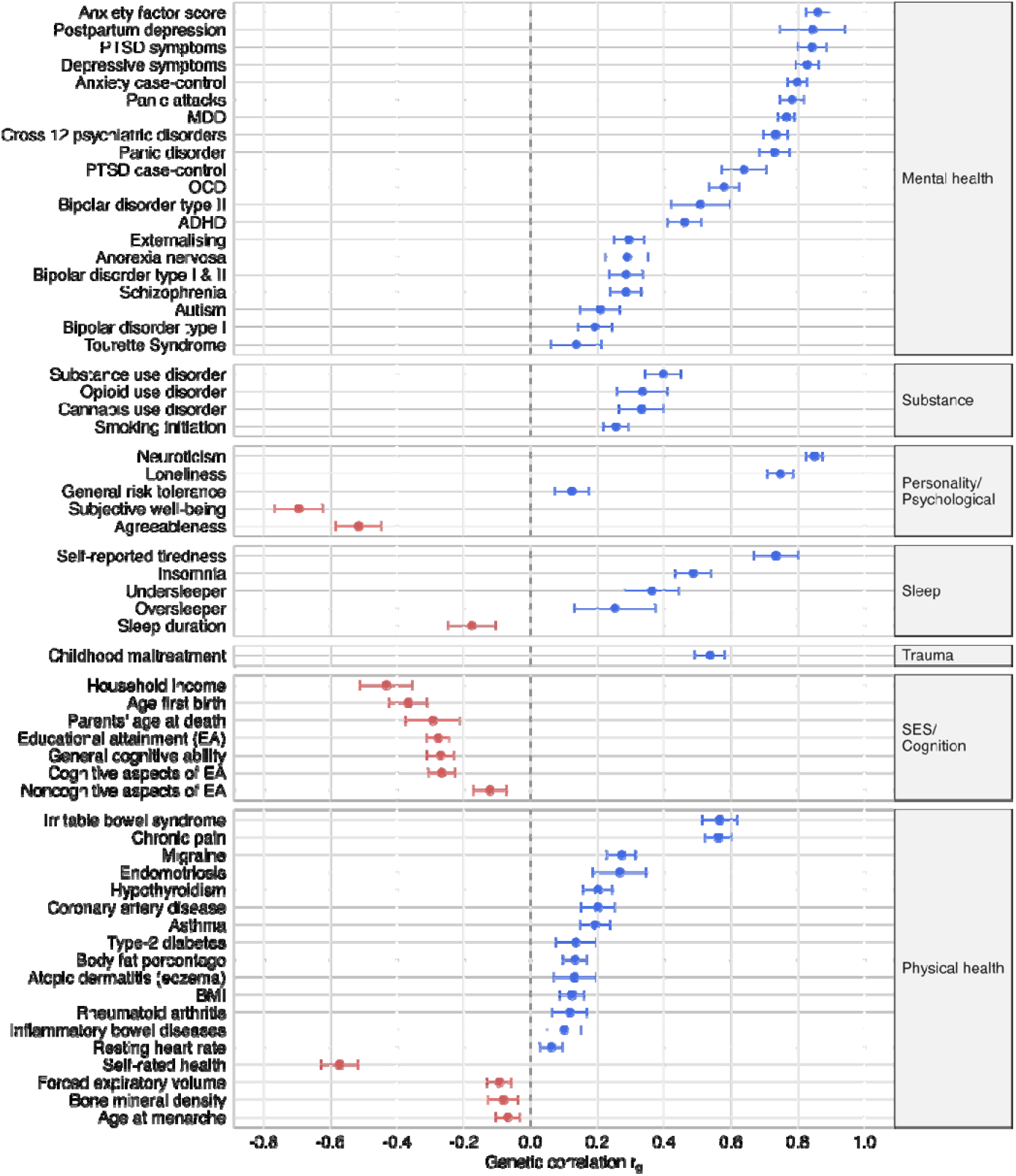
Genetic correlations between generalised anxiety symptom severity and a range of traits from existing genome-wide association studies, estimated with LDSC. All significant at a Bonferroni-corrected threshold of *p* < 4.76 ×0^−4^. Bars represent 95% confidence intervals. PTSD = post-traumatic stress disorder, MDD = major depressive disorder, OCD = obsessive-compulsive disorder, ADHD = attention deficit/hyperactivity disorder, BMI = adult body mass index, SES = socioeconomic status. Forced expiratory volume is a measure of lung function.

The strongest correlations were observed with quantitative internalising traits including neuroticism, depressive symptoms, and a genetic anxiety factor (r_g_ = 0.83 - 0.86), as well as with case-control phenotypes for anxiety and depression (r_g_ = 0.71 - 0.80). Moderate-to-strong estimates were also found for post-traumatic stress disorder (r_g_ = 0.64), insomnia (r_g_ = 0.49), irritable bowel syndrome (r_g_ = 0.57), and chronic pain (r_g_ = 0.56). Negative correlations were observed with socioeconomic status indicators including household income (r_g_ = −0.43). Smaller but significant correlations were identified with coronary artery disease, endometriosis, hypothyroidism, and migraine (r_g_ = 0.20 − 0.27). Most other associations with physical traits and illnesses were comparatively weaker and included positive correlations with resting heart rate, rheumatoid arthritis and atopic dermatitis, and a negative correlation with lung function (absolute r_g_ = 0.06 - 0.13).

### Conditional analyses

To assess whether genome-wide significant loci for generalised anxiety symptoms represented effects unique to this phenotype, we performed multi-trait conditional and joint analysis (mtCOJO^34^), conditioning on highly genetically correlated traits. Conditioning on neuroticism (67 instruments, GSMR b_xy_ = 0.47±0.02) resulted in 9 remaining genome-wide significant SNPs across 2 loci (Table S10). Conditioning on case-control anxiety (11 instruments, b_xy_ = 0.22±0.02) yielded 66 significant SNPs across 6 loci, and on depression (98 instruments, b_xy_ = 0.31±0.01) produced 76 SNPs across 6 loci. Approximately half of SNP effect sizes were attenuated, and standard errors had a median inflation of 7-9%, suggesting the loss of significance was not solely attributable to reduced power. Sign changes were observed in 17-20% of variants for each trait, consistent with instability as expected under high genetic correlations. The LDSC SNP-based heritability estimates of the conditioned summary statistics were significant (neuroticism h^2^ = 0.014, SE = 0.0010, *p* = 1.02×0^−42^; case-control anxiety h^2^ = 0.018, SE = 0.0011, *p* = 1.54×0^−59^; case-control depression h^2^ = 0.017, SE = 0.0011, *p* = 7.34×0^−51^), but block jack-knifing confirmed they were significantly lower than the original estimate (Z = 33.24, p = 3.3×0^−242^; Z = 28.61, p = 5.0×0^−180^; Z = 30.48, 4.24×0^−204^, respectively).

### Polygenic risk scores

To evaluate the within- and cross-ancestry generalisability of our GWAS findings, we generated a polygenic risk score (PRS) for GAD symptom severity using SBayesRC^35^ and tested its association with both quantitative and case-control anxiety outcomes in independent samples across ancestry groups (Table S11). The PRS significantly explained 2.9% of the variance (r^2^) in generalised anxiety symptom scores in an independent European ancestry sample (beta = 0.55, SE = 0.054, *p* = 8.9 × 10^−24^), 1.4% in an African ancestry sample (beta = 0.48, SE = 0.101, *p* = 2.9 × 10^−6^), and 1.2% in a South Asian ancestry sample (beta = 0.45, SE = 0.095, *p* = 2.0 × 10^−6^). For case-control anxiety, assuming 20% prevalence, the PRS explained 1.8% (beta = 0.25, SE = 0.057, *p* = 1.2 × 10^−5^) of the variance on the liability scale in a European ancestry sample, 1.7% (beta = 0.27, SE = 0.086, *p* = 2.0 × 10^−3^) in an African ancestry sample, and 1.4% (beta = 0.25, SE = 0.079, *p* = 1.3 × 10^−3^) in a South Asian ancestry sample. These R^2^ values represent the variance explained by the PRS beyond that accounted for by covariates.

### Positional and functional annotation

Functionally informed fine-mapping using PolyFun (v1.0.0)^36^ and SuSiE (v0.11.92)^37^ identified four putative causal variants with posterior inclusion probabilities (PIP) ≥ 0.95. Two of these were index variants for genome-wide significant loci: rs2392289 at locus 31 (PIP = 0.983) and rs72676302 at locus 58 (PIP = 0.978). The remaining two variants were in loci that did not reach genome-wide significance. One variant, rs72676302, had a high Combined Annotation Dependent Depletion (CADD) score of 19.78, suggesting a deleterious effect (above the suggested threshold of 12.37^38^), although there was little biological evidence that it is within a regulatory element (regulomeDB score = 5). Additionally, we identified 24 small credible causal sets (< 10 variants each) that cumulatively met the PIP threshold (Table S12).

SNP-level gene annotation was performed using FUMA (v1.6.5^39^) based on positional, expression quantitative trait loci (eQTL), and chromatin interaction (Hi-C) mappings (Table S13). We identified genes annotated by more than one method, with this convergence providing greater support for their involvement. This approach again highlighted *PCLO*, as well as *TMEM106B*, which has been associated with anxiety repeatedly^18,19,23^, and with depression^40^. In addition, multiple genes were identified that have previously been implicated in depression (e.g., *ERBB4, GRM7, VRK2, DCC, LRFN5, PCLO*) and schizophrenia (e.g., *ERBB4, VRK2*). Two genes, *SORCS3*^23^ and *MAD1L1*^17–19,26^, were implicated only by positional mapping in our analysis but have been associated with anxiety phenotypes in previous GWAS, lending further support to their relevance. While detailed functional information was limited for many of the mapped genes, several are posited to play roles in key neurotransmitter systems, including glutamatergic (*GRM7, HOMER1*), GABAergic (*ERBB4*), and dopaminergic (*DRD1*) signalling. In contrast, gene annotations for loci novel for associations with internalising traits did not indicate clear neurobiological pathways implicated in anxiety. For example, rs58179213 is proximal to multiple genes with evidence for involvement in diverse processes, including lung function (*SFTPC*), intestinal barrier function and immunoprotective inflammation (*FHIP2B*), and hair growth (*HR*). While such functions may influence anxiety via indirect pathways (e.g. gut-brain axis, somatic anxiety symptoms), replication of these loci is required to provide the greater power necessary to determine their relevance.

### Gene-based associations and enrichment

Gene-based association analysis was conducted using MAGMA^41^, which aggregates trait-SNP associations across all SNPs within a gene while accounting for LD. In total, 197 genes across 80 independent loci surpassed the Bonferroni-corrected significance threshold (*p* < 2.5 × 10^−6^; Table S14). The top associated gene was *PCLO* (*p* = 2.5 × 10^−21^), mirroring the SNP-based results.

To assess biological pathways significantly enriched for associations with GAD symptom severity, we performed pathway analysis in MAGMA with predefined gene sets (MsigDB v2023.1.Hs^42^; curated and gene ontology terms; Table S15). Six gene sets passed Bonferroni correction (*p* < 2.9 × 10^−6^): *postsynaptic membrane* (271 genes, *p* = 2.6 × 10^−8^), *synaptic membrane* (383 genes, *p* = 7.1 × 10^−8^), *axon* (627 genes, *p* = 5.3 × 10^−7^), *neurogenesis* (1627 genes, *p* = 8.6 × 10^−7^), *postsynapse* (648 genes, *p* = 9.2 × 10^−7^), and *generation of neurons* (1412 genes, *p* = 1.0 × 10^−6^). These results were unaffected by the exclusion of the MHC region, though the number of genes per set slightly decreased.

Gene-tissue expression analysis was conducted using catalogues of gene expression levels across different human tissues (Figs S2-5), with a Bonferroni correction applied within each tissue set. Among brain samples from 11 developmental stages (BrainSpan), only the two prenatal samples were significant (*p* = 6.3 × 10^−4^ and 2.5 × 10^−3^). In adult tissues (GTEx v8^43^), significant enrichment was observed in brain (*p* = 3.0 × 10^−9^) and pituitary (*p* = 4.9 × 10^−5^) tissues. Analysis of more specific tissue types revealed enrichment in 11 brain regions, most strongly in the frontal cortex, cortex, cerebellum, anterior cingulate cortex, and nucleus accumbens (all *p* < 9.3 × 10^−4^, Table S16).

### Drug targets

To explore therapeutic relevance, we ran DrugTargetor (v1.3^44^) to identify drug priorities with potential utility for clinical anxiety, based on their relevance for GAD symptom severity. We did not observe any significant enrichment of anxiety associations for the 1551 drug targets tested (Table S17). However, at the drug class level, we identified significant associations for Anatomical Therapeutic Chemical (ATC) classifications N06 ‘psychoanaleptics’ - including N06A antidepressants - N02A ‘opioids’, and N06B ‘psychostimulants’ (Bonferroni-adjusted *p* < 0.05; Table S18). With the exception of psychostimulants, these classes include drugs that have been used clinically to treat anxiety^45^. The enrichment for psychostimulants likely reflects shared dopaminergic and noradrenergic pathways involved in arousal and vigilance and underlying both anxiety and attention, consistent with pleiotropic genetic effects rather than direct therapeutic action.

## Discussion

In this genome-wide association meta-analysis of 693,869 individuals from 14 cohorts, we identified 80 genome-wide significant variants across 74 loci associated with quantitative measures of GAD symptom severity. This represents the largest number of genetic associations reported to date with anxiety symptoms. Approximately half of the identified loci replicated associations reported in previous anxiety GWAS^19–23,46^, while the remainder were novel.

The second strongest association was estimated for rs1476548 within *PCLO*, which was also implicated through positional mapping, eQTL mapping and gene-based association testing. *PCLO* encodes a protein involved in regulating presynaptic structure and neurotransmitter release. This gene has long been of interest in major depressive disorder (MDD^47^), with recent evidence also linking it to anxiety disorders^20,23^. Another gene of interest from our analysis was *SORCS3*, which was supported by multiple lines of evidence in the recent PGC-ANX case-control anxiety GWAS^23^. *SORCS3* plays a role in postsynaptic functioning and glutamate receptor regulation, particularly in the hippocampus^48^. It has been linked to memory and learning processes, specifically synaptic depression and fear extinction^49^, as well as mental health and neurodevelopmental conditions including MDD, Tourette syndrome, attention-deficit/hyperactivity disorder, and autism^50,51^.

Our SNP-based heritability estimate (5.9%) aligned with an existing GWAS of GAD symptom severity (5.6%)^26^ while remaining lower than liability scale estimates from case-control anxiety meta-analyses^23^. In contrast to traditional case-control phenotyping, which aims to maximise clinical specificity through diagnostic thresholds, our approach leveraged the full spectrum of symptom variability, increasing power for discovery^52^ and capturing genetic risk relevant to both subclinical and clinical presentations. The key differences between clinical diagnoses and symptom severity measures relate to the presence of distress and impairment, and symptom duration. While subclinical symptoms can still cause distress and impairment, this is not always true for lower levels of anxiety severity, potentially contributing to some diagnosis-specific genetic variance. Similarly, diagnostic tools often assess lifetime occurrence and require symptoms to be present for a minimum period of time (six months for GAD), whereas symptom severity scales typically capture recent experiences (e.g., past two weeks), introducing greater susceptibility to transient fluctuations and measurement noise. Consistent with this, GWAS of depression symptom severity typically yield lower SNP-based heritability estimates than case-control analyses^53–55^. We aimed to partially address and control for temporal fluctuations and better approximate a stable underlying trait^16^ by incorporating assessments from multiple timepoints into our analysis, where available. In addition, our meta-analysis combined data from multiple cohorts with differences in phenotype definitions, genotyping arrays, imputation methods, quality control procedures, and population structure adjustments, which may have introduced further heterogeneity and reduced the observed SNP-based heritability. By contrast, single-cohort studies with individual-level data are more homogeneous and can implement alternatives to summary-statistics based methods. Despite our lower heritability estimate, the strong genetic correlation observed between our phenotype and case-control anxiety suggests that GAD symptom severity captures much of the same genetic risk. This finding is consistent with a recent analysis of obsessive-compulsive symptoms^56^. Quantitative, symptom-based approaches may be particularly well-suited to genetic studies of anxiety, given the high burden of anxiety symptoms observed across other mental health conditions^8–11^. In this context, efforts to isolate ‘pure’ anxiety cases may be both methodologically challenging and reflect an unusual clinical phenotype that is invalid for most individuals with anxiety.

There was a broad range of significant genetic correlations across both mental and physical health conditions, mirroring the frequent co-occurrence with anxiety symptoms and widespread pleiotropy. A strong genetic correlation was observed with neuroticism, a well-established risk factor for anxiety^56^. This likely reflects both genuine shared liability and conceptual or item-level overlap between the measures. Conditional analyses indicated extensive overlap between loci associated with generalised anxiety symptoms and those implicated in neuroticism, case-control anxiety and depression. Many genome-wide significant loci may therefore reflect a broader neuroticism-related liability, although the presence of anxiety-specific effects cannot be excluded due to the statistical noise introduced by conditioning on highly genetically correlated traits. Many of the genetic correlations aligned with findings from a genomicSEM analysis of anxiety symptoms and disorder^22^, including strong associations with irritable bowel syndrome and chronic pain, and a moderate association with migraine. These results do not necessarily reflect horizontal pleiotropy, potentially arising from the experience of these conditions eliciting uncertainty and worry that contribute to anxiety.

Polygenic scores derived from our genome-wide association meta-analysis demonstrated within- and cross-ancestry generalisability, significantly explaining 1.2% to 2.9% of the variance in GAD symptom severity in European, African, and South Asian ancestry samples. Across these samples, the PRS also accounted for 1.4% to 1.8% of the variance in case-control anxiety on the liability scale. While broadly consistent with the 0.5% to 2.3% range reported in the PGC-ANX case-control analysis^23^, direct comparisons are limited by methodological differences in PRS construction and target sample composition. A key limitation of the present meta-analysis is its restriction to cohorts of European ancestry, due to lack of data for other ancestries at sufficient scale for GWAS analysis. Our PRS findings support a degree of shared genetic influence with African and South Asian populations. Ancestry-specific modelling across diverse populations remains necessary to identify population-specific risk loci, such as that identified in a previous analysis of African-American participants^26^, and ensure equitable benefits from genetic discoveries. Overall, these findings provide additional evidence that quantitative phenotyping can effectively capture genetic signal relevant to clinical anxiety.

The GAD symptom severity measures and ascertainment methods varied across contributing cohorts, although most assessed symptoms using the GAD-7. While widely adopted across clinical and research contexts, the GAD-7 does not comprehensively assess all DSM-5 GAD symptoms - omitting sleep and concentration problems - and is not designed to capture symptoms of fear-based anxiety (i.e. phobias, social anxiety disorder) or panic disorder. This limits the generalisability of our findings across anxiety disorders, particularly in light of evidence for partially distinct phenotypic and genetic contributions to GAD compared with fear-based disorders^57,58^. Expanding future studies to incorporate a broader range of anxiety symptom measures will enable more robust, transdiagnostic translation of findings. Furthermore, future work could examine the genetic architecture of individual symptom domains, such as cognitive versus physiological symptoms, to better understand the biological specificity of these. Our subgroup analyses based on measure and ascertainment method were largely underpowered to reliably estimate SNP-based heritability or correlations. Although sufficiently powered comparisons indicated high genetic overlap, we cannot be certain that all cohorts captured the same underlying genetic architecture. While population-based cohorts allow assessment of the full range of symptoms, symptom severity measures typically better distinguish variation at the upper end of the distribution. This results in highly skewed symptom severity scores, as most participants report few or no symptoms, whereas individuals in clinical cohorts typically report more symptoms. Combining population-based cohorts with studies selecting on diagnosis introduces some heterogeneity and may limit generalisability to broader populations, but can help yield a more normally distributed phenotype for GWAS analysis. This combination may have improved statistical power for detecting associations in our study.

Our quantitative GWAS of GAD symptom severity identified more genome-wide significant loci than a slightly larger and mostly overlapping case-control anxiety study (N = 852,222; 122,341 cases)^23^, with many loci replicated across the two methods. This aligns with expectations under the liability-threshold model when considering common conditions such as anxiety, whereby quantitative traits generally offer greater statistical power than case-control designs of equal sample size^52^. Beyond identifying the largest number of anxiety-associated loci to date, our results implicate key neurobiological pathways, including synaptic function and neurotransmission, and notable genes such as *PCLO* and *SORCS3*. These findings demonstrate that a quantitative anxiety symptom-based phenotype can reveal biologically meaningful signals and complements insights from case-control designs. Clinically ascertained samples remain essential for identifying disorder-specific biology and mapping genetic risk to diagnostic presentations, however, obtaining clinical cases at sufficient scale for binary genome-wide analyses is challenging. Although electronic health records offer an efficient option, these are limited to individuals seeking and receiving medical attention. Quantitative, symptom-based approaches within biobanks and population studies therefore offer a promising scalable alternative for advancing the field of anxiety genetics. Moving forward, the combination of these with deeply phenotyped clinical cohorts will be crucial for translating genetic insights into diagnostic and therapeutic advances. Together, these approaches can elucidate the biological continuum of anxiety, from healthy stress responses to debilitating disorder. Given the high and rising rates of anxiety, especially in young adults, it is more important than ever to improve our ability to identify and understand sources of risk. Despite its public health impact, progress in anxiety genetics lags behind other major mental health conditions. We hope our findings encourage a new wave of genome-wide investigations leveraging existing but potentially underutilised anxiety severity data in genotyped cohorts, accelerating our progress in understanding the genetic architecture of anxiety.

## Methods

### Participants and measures

We meta-analysed data from 14 international cohorts (N = 693,869); 13 PGC-ANX studies with generalised anxiety symptom data, and summary statistics from a pre-existing GWAS^26^. We performed a meta-analysis with access to individual participant data^59^, such that each PGC-ANX cohort performed genome-wide association analyses specifically for this study and shared summary statistics with the core analytical team. The majority of the sample (70%) had completed the GAD-7 or closely related brief self-report measures assessing recent anxiety symptoms. The remaining 30% used other brief self-report anxiety scales (Table S1), each available in at least 3,000 individuals. We analysed total sum scores, with higher scores indicating greater severity of symptoms. If participants were missing data on <25% of measure items, the missing scores were imputed with the participant’s mean score of the other items. Participants with ≥25% missing data were excluded from analysis. Several cohorts had assessed anxiety symptoms on two or more occasions. Longitudinal twin studies have shown that symptom stability is primarily driven by genetic factors^16,60,61^ and stability extracted from repeated assessments yields higher heritability estimates than single timepoints^16^. For cohorts with anxiety assessments from three or more timepoints (12% of the sample), a latent factor was created in R with the package *lavaan*^62^, the *predict* function and an ML estimator. For cohorts with two timepoints (44%), a mean score was calculated. All scores, whether single timepoint, mean or factor score, were standardised to have a mean of zero and a standard deviation of one. Given the high comorbidity of anxiety and other mental health conditions, no additional exclusions were applied beyond those defined by each study. For two cohorts - GLAD+ and the UK Biobank - individual-level data were merged prior to the GWAS. Participants from clinical cohorts had been recruited based on a lifetime history of depression or anxiety, as assessed by self-reported diagnostic questionnaires.

### Meta-analysis

Table S3 provides details of the studies that contributed to this meta-analysis, which were: Australian Genetics of Depression Study (AGDS)^63^, Avon Longitudinal Study of Parents and Children (ALSPAC)^64,65^, CoLaus|PsyCoLaus^66^, Estonian Biobank^67^, Generation Scotland^68^, NIHR Bioresource Genetic Links to Anxiety and Depression Study (GLAD+^69^), Lifelines^70^, MEGA TRR58^71^, Million Veteran Program^26^, Norwegian Mother, Father, and Child Cohort Study^72^, Providing Tools for Effective Care and Treatment of Anxiety Disorders (PROTECT-AD^73^), Twins Early Development Study (TEDS)^74^, Tracking Adolescents’ Individual Lives’ Survey (TRAILS^75^), and UK Biobank^76^. Each cohort performed genotyping using microarray platforms and imputed genotypes using ancestry matched panels, primarily Haplotype Reference Consortium^77^. Rigorous quality control procedures were applied, including filters on sample and variant call rates, sex concordance, and excessive heterozygosity (full details in Table S3). The one set of pre-existing summary statistics was from an analysis in the MVP, obtained through the database of Genotypes and Phenotypes (dbGaP; phs001672). Most groups adopted a mixed linear model approach and retained related individuals in their GWAS. Where applicable, covariates such as ancestry principal components and genotyping batch were included. We did not include age or sex as covariates, as they are not confounders of genetic effects and may represent effect moderators of interest warranting dedicated follow-up investigation, rather than variables to be adjusted for. All resulting summary statistics were on the GRCh37 genome assembly (b37/Hg19). Prior to meta-analysis, variant-level quality control was performed across the summary statistics, excluding those with minor allele frequency (MAF) <1% or imputation accuracy score <0.6. Variants present in fewer than half the contributing cohorts were excluded, resulting in a total of 7,499,431 autosomal SNPs. X-chromosome data were analysed from seven cohorts (166,852 variants), with male genotypes coded as diploid (0/2) and sex included as a covariate. The meta-analysis was conducted in METAL^78^ using an inverse-variance weighted, standard-error based approach. The beta values from the meta-analysis represent the associated change in standard deviation units of generalised anxiety symptom score per additional copy of the effect allele.

Heterogeneity across cohorts was assessed by inspecting the p-values from Cochran’s Q-test as implemented in METAL. In addition, we calculated the median I^2^ values (HetISq in METAL) for SNPs reaching genome-wide significance (*p* < 5×0^−8^) and for independent lead SNPs. We also estimated genetic correlations between contributing cohorts using LDSC^33^. The inclusion of clinical alongside community-based cohorts offered greater representation across the full range of the anxiety symptom severity, increasing statistical power as evidenced in a recent depression GWAS^80^. However, due to the risk of bias or confounding from differences in study design and phenotyping, we performed subgroup meta-analyses stratified by anxiety measure and excluding clinical cohorts (Table S2). Meta-analyses of the measure and ascertainment subgroups were performed in METAL, and genetic correlations between the groups estimated using LDSC^33^. For both cohort and subgroup genetic correlations, most pairwise comparisons were not sufficiently powered (i.e. heritability z-scores < 4 for one or both sets of sumstats^79^) to draw conclusions.

To identify LD independent significant SNPs and loci, clumping was performed in FUMA^39^ (v1.6.5). The r^2^ threshold for independent significant SNPs was 0.1, and for lead SNPs was 0.05, within a 500kb window. Genome-wide significance was defined using the conventional threshold (*p* < 5×0^−8^).

To determine the novelty of our results, we cross-referenced significant loci with published trait associations from the GWAS Catalog^81^ using LDTrait^82^, applying an r^2^ threshold of > 0.1 and a 500kb window. Novelty was strictly defined as having no prior associations with internalising traits including anxiety, depression, neuroticism, and worry. To supplement this, we compared our results with recent anxiety and depression studies not yet available in the GWAS Catalog^20,22,23,80^. Overlapping significant loci were identified with BEDtools^83^ and LD assessed using a threshold of r^2^ > 0.1. The investigation of novelty also revealed the extent to which our results replicated previous findings. We additionally determined novelty specifically for anxiety, whether assessed as symptom severity or a case-control phenotype. Many of the 14 cohorts in our meta-analysis overlap with prior case-control anxiety meta-analyses, with the exception of GLAD+, Lifelines, ProtectAD, TEDS, and MEGA (approximate N = 110,000). In some instances, the cohort sample composition differs due to the availability of quantitative versus diagnostic information.

### SNP-based heritability and genetic correlations with external traits

We estimated SNP-based heritability via SBayesRC^35^. This provided an estimate of the proportion of variance in quantitative anxiety attributable to variation in the common SNPs present in this meta-analysis. We inspected the LDSC^32^ genomic inflation factor (λ_GC_) and intercept to evaluate the contribution of potential confounding relative to polygenicity. Genetic correlations were also computed using LDSC with 105 GWAS summary statistics covering a broad range of phenotypes and applying a Bonferroni-corrected p-value threshold of 4.76×0^−4^. High genetic correlations, such as those often observed between anxiety, depression, and neuroticism, do not necessarily indicate identical biology; even when most loci are shared, traits may involve different biological pathways, tissue enrichments, or show individual patterns of relationships with other traits. Identifying unique genetic influences on anxiety is important to better understand its specific aetiology and inform potential treatment pathways. However, conditional analyses, especially in the presence of strong genetic correlations, are statistically challenging and require substantial power, and therefore should be interpreted cautiously. We ran mtCOJO^34^, which performs conditional analyses between summary statistics to provide marginal effect estimates for the trait of interest. We conditioned our anxiety meta-analysis on depression diagnosis^80^ (98 index SNPs), neuroticism^84^ (67 index SNPs) and anxiety diagnosis^23^ (11 index SNPs). Depression symptoms^85^ was underpowered (2 index SNPs) for the analysis. We also estimated the SNP-based heritability of each set of conditioned summary statistics with LDSC and used block jack-knifing to compare these to the unconditioned heritability estimate.

### Polygenic risk scores

To evaluate the within and cross-ancestry validity of our GWAS, we calculated GAD symptom severity polygenic risk scores (PRS) in independent samples from the UK Biobank^76^ and Prospective Imaging Study of Ageing (PISA)^86^. We then performed regressions between our PRS and quantitative anxiety, using GAD-7 scores, as well as case-control anxiety, as defined by a self-reported diagnostic questionnaire or self-report of a diagnosis from a health professional. Specifically, we used SBayesRC^35^ to calculate PRS in European, African, and South-Asian ancestry samples, excluding related individuals. SBayesRC is a Bayesian regression method that uses GWAS summary statistics to estimate SNP effect sizes while accounting for LD and polygenic architecture. It extends the SBayesR framework by incorporating functional annotations or prior biological information, improving the detection of likely causal variants and enhancing predictive accuracy for complex traits. We conducted linear regressions to assess the variance explained in GAD symptom severity by the PRS in each sample (European N = 3,452; African N = 1,581; South Asian N = 1,813). For case-control status, we performed logistic regressions to estimate effects in the target samples European total n = 3,107, case n = 407; African total n = 1,303, case n = 218; South Asian total n = 1,549, case n = 265). We calculated Nagelkerke’s r^2^ for our PRS on the liability scale using the observed linear regression r^2^ and corresponding formula^87^, assuming a population prevalence of 20%. All regressions included the first 10 ancestry-specific PCs and genotyping batch as covariates. The variance explained by the PRS was calculated as the difference in R^2^ between a full model including the PRS and covariates, and a null model with only covariates.

### Positional and functional annotation

We used PolyFun^36^ to estimate per-SNP heritabilities, leveraging a regularised extension of stratified-LDSC (s-LDSC) applied to the v.2.2.UKB baseline-LF model annotations, which captures heritability enrichment related to allele frequency, LD and variant function. These prior causal estimates were then used for fine-mapping in SuSiE^37^, limiting to a maximum of one causal SNP per locus. We extracted annotations at a Posterior Inclusion Probability (PIP) threshold of ≥ 0.95 and created credible causal sets containing the minimum set of ranked variants that cumulatively met this threshold. Unlike standard definitions of credible causal sets in SuSiE, we did not require a minimum pairwise r^2^ between variants in a set, as the PolyFun + SuSiE pipeline does not incorporate LD estimates.

We performed SNP-level gene annotation using FUMA^39^ (v1.6.5), integrating three complementary methods: positional mapping (based on physical proximity to genes), expression quantitative trait loci (eQTL) mapping (linking variants to gene expression), and chromatin interaction mapping (using Hi-C data to identify regulatory interactions). eQTL mapping used significant SNP-gene pairs and eQTLs from the brain tissue datasets GTEx v8 Brain^43^ (13 regions) and BRAINEAC^88^ (10 regions), and average expressions across these, applying a false discovery rate (FDR) threshold of *p* < 0.05. Chromatin interaction mapping employed Hi-C brain tissue data (dorso-lateral prefrontal cortex, hippocampus, left and right ventricles)^89^ and adult and foetal cortex^90^, with an FDR threshold of *p* < 1 × 10^−6^. These methods differ in their underlying biological rationale and may implicate different genes. Genes identified by two or more mapping approaches were therefore highlighted, as convergence across the methods increased our confidence in the potential functional relevance of a gene.

### Gene-based associations and enrichment

Gene-based association, gene-set, and gene-tissue expression enrichment analyses were performed in MAGMA^41^ (v1.08) via FUMA^39^ (v1.6.5). These analyses aimed to identify genes associated with GAD symptom severity, biological pathways enriched for associated genes, and relevant tissues where genes are preferentially expressed, offering insight into the potential biological mechanisms underlying our findings. For gene-based associations, we tested 19,954 genes, applying a Bonferroni-corrected significance threshold of *p* < 2.51 × 10^−6^. SNPs were mapped to genes using a 35kb upstream and 10kb downstream window. Gene-set analyses were performed using 6,494 curated gene sets (‘c2.all’) and 10,529 gene-ontology (GO) terms (‘c5.bp’, ‘c5.cc’ and ‘c5.mf’) from the Molecular Signatures Database (MSigDB^42^; v2023.1.Hs). Significance was determined by a Bonferroni-corrected threshold of *p* < 2.94 × 10^−6^. For tissue enrichment we tested relationships between trait-associated genes and gene expression in human tissues, using data from BrainSpan (brain samples from 11 general developmental stages and 29 specified ages) and GTEx v8 (covering 30 general and 54 specific tissue types).

### Drug targets

We examined whether genes associated with GAD symptom severity were associated with individual drugs and drug classes using the DrugTargetor^44^ method (November 2020 update). DrugTargetor integrates MAGMA gene-level association results with curated drug-gene interaction databases (ChEMBL^91,92^ and DGIdb^93^). We used MAGMA (v1.10) to prioritise associated genes within windows 35kb upstream and 10kb downstream. We hypothesised drug action within the nervous system, a maximum of 1,551 unique drugs and 163 drug classes. To assess the enrichment of drug classes we calculated the area under the enrichment curve (AUC), where 50% indicates random enrichment and 100% optimal enrichment, and AUC significance was assessed using one-sided Wilcoxon-Mann-Whitney tests.

## Supporting information

Supplementary Figuress

## Data and code availability

Summary statistics will be made available on the PGC data-download page (https://pgc.unc.edu/for-researchers/download-result). Analytical code is available via Github https://github.com/megskelton/gad-sympt-metagwas.

## Acknowledgements

**AGDS:** We are indebted to the participants for giving their time to contribute to this study. We thank all the people who helped in the conception, implementation, beta testing, media campaign, and data cleaning. The Australian Genetics of Depression Study was funded by grant 108663 from the Australian National Health and Medical Research Council (NHMRC). This work was supported by NHMRC Investigator Grants to BLM (2017176); NRW (1173790); NGM (1172990); SEM (1172917 and 2025674) and IBH (2016346). EMB received funding from the PRE-EMPT NHMRC Centre for Research Excellence (1198304) and the University of Queensland Health Research Accelerator Program.

**ALSPAC:** We are extremely grateful to all the families who took part in this study, the midwives for their help recruiting them, and the whole ALSPAC team, which includes interviewers, computer and laboratory technicians, clerical workers, research scientists, volunteers, managers, receptionists and nurses. The UK Medical Research Council and Wellcome (Grant ref: 217065/Z/19/Z) and the University of Bristol provide core support for ALSPAC. This publication is the work of the authors and ASFK will serve as guarantor for the ALSPAC contents of this paper. Genome-wide genotyping data was generated by Sample Logistics and Genotyping Facilities at Wellcome Sanger Institute and LabCorp (Laboratory Corporation of America) using support from 23andMe. Ethical approval for the study was obtained from the ALSPAC Ethics and Law Committee and the Local Research Ethics Committees.

**CoLaus**|**PsyCoLaus:** The CoLaus|PsyCoLaus study was supported by unrestricted research grants from GlaxoSmithKline, the Faculty of Biology and Medicine of Lausanne, the Swiss National Science Foundation (grants 3200B0–105993, 3200B0-118308, 33CSCO-122661, 33CS30-139468, 33CS30-148401, 33CS30_177535, 3247730_204523 and 320030_220190) and the Swiss Personalized Health Network (grant 2018DRI01).

**Estonian Biobank:** We acknowledge the Estonian Biobank participants and the Estonian Biobank Research Team (Andres Metspalu, Lili Milani, Tõnu Esko, Reedik Mägi, Mait Metspalu, Mari Nelis, Georgi Hudjashov). Data analysis was carried out in part in the High-Performance Computing Center of University of Tartu. This research was supported by the Estonian Research Council (grant PSG615) and the Estonian Centre of Excellence for Well-Being Sciences, funded by grant TK218 from the Estonian Ministry of Education and Research. The research was conducted using the Estonian Center of Genomics/Roadmap II funded by the Estonian Research Council (project number TT17).

**Generation Scotland:** We would like to thank the participants and staff of Generation Scotland. The work presented is supported by Wellcome Trust (226770/Z/22/Z, 220857/Z/20/Z, 216767/Z/19/Z) and UK Research and Innovation (MR/Z50354X/1, MR/Z000548/1). Genotyping of GS samples was funded by the MRC and Wellcome Trust (104036/Z/14/Z). GS also received support from the Chief Scientist Office of the Scottish Government Health Directorates (CZD/16/6) and the Scottish Funding Council (HR03006) GLAD+ (including NIHR BioResource): We thank NIHR BioResource volunteers for their participation, and gratefully acknowledge NIHR BioResource centres, NHS Trusts and staff for their contribution. We thank the National Institute for Health and Care Research, NHS Blood and Transplant, and Health Data Research UK as part of the Digital Innovation Hub Programme. The views expressed are those of the author(s) and not necessarily those of the NHS, the NIHR or the Department of Health and Social Care. We gratefully acknowledge the participation of the NIHR BioResource Centre Maudsley, Biomedical Research Centre at South London and Maudsley NHS Foundation Trust and King’s College London volunteers and thank the staff for their help with volunteer recruitment. We thank the NIHR Biomedical Research Centre at South London and the Maudsley NHS Foundation Trust and King’s College London for funding. This study represents independent research supported by the NIHR Biomedical Research Centre BioResource at South London and Maudsley NHS Foundation Trust and King’s College London. We gratefully acknowledge capital equipment funding from the Maudsley Charity (Grant Ref. 980) and Guy’s and St Thomas’s Charity (Grant Ref. STR130505).

**Lifelines Cohort Study:** The Lifelines Biobank initiative has been made possible by funding from the Dutch Ministry of Health, Welfare and Sport, the Dutch Ministry of Economic Affairs, the University Medical Center Groningen (UMCG the Netherlands), University of Groningen and the Northern Provinces of the Netherlands. The generation and management of GWAS genotype data for the Lifelines Cohort Study is supported by the UMCG Genetics Lifelines Initiative (UGLI). UGLI is partly supported by a Spinoza Grant from NWO, awarded to Cisca Wijmenga. The authors wish to acknowledge the services of the Lifelines Cohort Study, the contributing research centers delivering data to Lifelines, and all the study participants.

**MEGA TRR58:** This project was funded by the German Research Foundation (DFG) – project no. 44541416 TRR 58 ‘Fear, Anxiety, Anxiety Disorders’, project Z02/1-3 and project no. 499262975. This work was supported by the German Research Foundation (DFG, grant FOR2107 DA1151/5-1, DA1151/5-2, DA1151/9-1, DA1151/10-1, DA1151/11-1 to UD; SFB/TRR 393, project grant no 521379614) and the Interdisciplinary Center for Clinical Research (IZKF) of the medical faculty of Münster (grant Dan 3/022/22 to UD). We would like to thank Manuel Kuhn for help with data acquisition.

**MoBa:** We are grateful to all the participating families in Norway who take part in this on-going cohort study. For generating high-quality genomic data, we thank the Norwegian Institute of Public Health (NIPH), the HARVEST collaboration (funded by the Research Council of Norway #229624), the NORMENT Centre at the University of Oslo (funded by the Research Council of Norway #223273), the Center for Diabetes Research at the University of Bergen (funded by the ERC AdG project SELECTionPREDISPOSED), deCODE Genetics, the Research Council of Norway, the SouthEastern and Western Norway Regional Health Authorities, the ERC AdG, Stiftelsen KG Jebsen, the Trond Mohn Foundation, and the Novo Nordisk Foundation. The MoBa analysis was performed on the Tjeneste for Sensitive Data (TSD) facilities, owned by the University of Oslo, operated and developed by the TSD service group at the University of Oslo, IT-Department (USIT), using resources provided by Sigma2—the National Infrastructure for High Performance Computing and Data Storage in Norway (UNINETT). Funding from the Research Council of Norway (RCN #324620, #336085), Helse Sør-Øst (#2020022) and NordForsk (#156298). ECC is supported by funding from the Research Council of Norway (#274611) and the South-Eastern Norway Regional Health Authority (#2021045). ECC is a member of the MRC Integrative Epidemiology Unit at the University of Bristol which is supported by the Medical Research Council and the University of Bristol (MC_UU_00032/1).

**MVP:** The authors thank Million Veteran Program (MVP) staff, researchers, and volunteers, who have contributed to MVP, and especially participants who previously served their country in the military and now generously agreed to enroll in the study. (See https://www.research.va.gov/mvp/ for more details). The citation for MVP is Gaziano, J.M. et al. Million Veteran Program: A mega-biobank to study genetic influences on health and disease. J Clin Epidemiol 70, 214-23 (2016). This research is based on data from the Million Veteran Program, Office of Research and Development, Veterans Health Administration.

**PISA:** The Prospective Imaging Study of Ageing (PISA) was funded by the National Health and Medical Research Council (NHMRC) of Australia [Grant ID: APP1095227].

**PROTECT-AD** (Providing Tools for Effective Care and Treatment of Anxiety Disorders): is one out of nine research consortia in the German federal research programme “Research Network on Mental Disorders”, funded by the Federal Ministry of Education and Research (BMBF; www.fzpe.de), grant number: 01EE1402. A complete list of project publications can be found at www.fzpe.de. Recruitment of Protect-AD was funded by the BMBF (Protect-AD, project P5, 01EE1402F).

**TEDS:** We gratefully acknowledge the ongoing contribution of the Twins Early Development Study (TEDS) participants and their families. TEDS is funded by a UK Medical Research Council (MRC) programme grant (MR/V012878/1) to TC Eley (previously MR/M021475/1 to R Plomin).

**TRAILS:** This research is part of the TRacking Adolescents’ Individual Lives Survey (TRAILS). Participating centers of TRAILS include various departments of the University Medical Center and University of Groningen, the University of Utrecht, and the Parnassia Psychiatric Institute, all in the Netherlands. TRAILS has been financially supported by grants from the Netherlands Organization for Scientific Research NWO (Medical Research Council program grant GB-MW 940-38-011; ZonMW Brainpower grant 100-001-004; ZonMw Risk Behavior and Dependence grant 60-60600-97-118; ZonMw Culture and Health grant 261-98-710; Social Sciences Council medium-sized investment grants GB-MaGW 480-01-006 and GB-MaGW 480-07-001; Social Sciences Council project grants GB-MaGW 452-04-314 and GB-MaGW 452-06-004; ZonMw Longitudinal Cohort Research on Early Detection and Treatment in Mental Health Care grant 636340002, NWO large-sized investment grant 175.010.2003.005; NWO Longitudinal Survey and Panel Funding 481-08-013 and 481-11-001; NWO Vici 016.130.002, 453-16-007/2735, and Vi.C 191.021; NWO Gravitation 024.001.003), the Dutch Ministry of Justice (WODC), the European Science Foundation (EuroSTRESS project FP-006), the European Research Council (ERC-2017-STG-757364 en ERC-CoG-2015-681466), Biobanking and Biomolecular Resources Research Infrastructure BBMRI-NL (CP 32), The Gratema foundation, the Jan Dekker foundation, the participating universities, and Accare Centre for Child and Adolescent Psychiatry. Statistical analyses were carried out on the Genetic Cluster Computer (http://www.geneticcluster.org), which is financially supported by the Netherlands Scientific Organization (NWO 480-05-003) along with a supplement from the Dutch Brain Foundation. We are grateful to everyone who participated in this research or worked on this project to make it possible.

**UK Biobank:** This research has been conducted using the UK Biobank Resource under Application Number 82087. This work uses data provided by patients and collected by the NHS as part of their care and support.

**Additional:** The authors acknowledge the use of the King’s College London research computing facility, CREATE (https://doi.org/10.18742/rnvf-m076). NS received funding from a UKRI Future Leaders Fellowship [grant number MR/T04327X/1] and the UK Dementia Research Institute award number UK DRI-5008 through UK DRI Ltd. NM acknowledges funding from the NHMRC. OAA received EU, RCN, NIH, NordForsk, SouthEast Health Authority funding. ASFK is supported by a Wellcome Early Career Award (Grant ref: 227063/Z/23/Z). EC is supported by the RCN (#274611) and HSØ (#2021045). LJH was supported by funding from the Norwegian South-East Regional Health Authority (#s 2021045, 2019097, 2022083). AH acknowledges the South-Eastern Norway Regional Health Authority (#2020022) and the Research Council of Norway (#274611; #336085). JH acknowledges NIMH 5R01MH124847. SM acknowledges funding from NHMRC APP1172917 and APP2025674. BLM was supported by an NHMRC Investigator Grant (APP2017176).

**23andme:** This paper uses summary statistics from analyses with 23andme participants to perform genetic correlations. We would like to thank the research participants and employees of 23andMe, Inc. for making this work possible.

## Declaration of interests

Prof Breen has received honoraria, research or conference grants and consulting fees from Illumina, Otsuka, and COMPASS Pathfinder Ltd.

Prof Ole A Andreassen - Consultant Precision Health and Cortechs.ai, speaker’s honorarium from Lundbeck, Otsuka, Janssen, and Lilly.

Prof Katharina Domschke is a member of the Neurotorium Editorial Board of the Lundbeck Foundation and has received speaker’s honoraria by Janssen Cilag GmbH, not related to the subject of this manuscript.

Prof Hickie is a Professor of Psychiatry and the Co-Director of Health and Policy, Brain and Mind Centre, University of Sydney. He has led major public health and health service development in Australia, particularly focusing on early intervention for young people with depression, suicidal thoughts and behaviours and complex mood disorders. He is active in the development through codesign, implementation and continuous evaluation of new health information and personal monitoring technologies to drive highly-personalised and measurement-based care. He holds a 3.2% equity share in Innowell Pty Ltd that is focused on digital transformation of mental health services.

All other authors declare no conflict of interests.

## Consortia

**Anxiety Working Group of the Psychiatric Genomics Consortium (PGC-ANX):** Strom, Nora, I. 1, 2, 3, 4; Verhulst, Brad 5; Bacanu, Silviu-Alin 6; Cheesman, Rosa 7; Purves, Kirstin, L. 8; Gedik, Hüseyin 9, 10, 11; Mitchell, Brittany, L. 12, 13; Kwong, Alex, S. 14, 15; Faucon, Annika, B. 16; Singh, Kritika 17, 18; Medland, Sarah 12; Colodro-Conde, Lucia 12, 19; Krebs, Kristi 20; Hoffmann, Per 21, 22; Herms, Stefan 21, 23, 22; Gehlen, Jan 24; Ripke, Stephan 25, 26; Awasthi, Swapnil 25; Palviainen, Teemu 27; Tasanko, Elisa, M. 28; Peterson, Roseann, E. 9, 6; Adkins, Daniel, E. 29; Shabalin, Andrey, A. 29; Adams, Mark, J. 30; Iveson, Matthew, H. 30; Campbell, Archie 31; Thomas, Laurent, F. 32, 33, 34, 35; Winsvold, Bendik, S. 36, 37, 38; Drange, Ole Kristian 39, 40, 41, 42, 43; Børte, Sigrid 44, 45, 37; ter Kuile, Abigail, R. 8, 46, 47; Nguyen, Tan-Hoang 11; Meier, Sandra, M. 48; Corfield, Elizabeth, C. 49, 50; Hannigan, Laurie 51, 49, 52; Levey, Daniel, F. 53, 54; Czamara, Darina 55; Weber, Heike 56; Choi, Karmel, W. 57, 58; Pistis, Giorgio 59; Couvy-Duchesne, Baptiste 12, 60, 61; Van der Auwera, Sandra 62; Teumer, Alexander 63, 62; Karlsson, Robert 64; Garcia-Argibay, Miguel 65, 64; Lee, Donghyung 66; Wang, Rujia 67; Bjerkeset, Ottar 68, 39; Stordal, Eystein 69, 39; Bäckmann, Julia 3; Salum, Giovanni, A. 70, 71; Zai, Clement, C. 72, 73, 74, 75, 76; Kennedy, James, L. 72, 73, 74; Zai, Gwyneth 72, 73, 74; Tiwari, Arun, K. 72, 73, 74; Heilmann-Heimbach, Stefanie 21; Schmidt, Börge 77; Kaprio, Jaakko 27; Kennedy, Martin, M. 78; Boden, Joseph 79; Havdahl, Alexandra 49, 51, 7, 14; Middeldorp, Christel, M. 80, 81; Lopes, Fabiana, L. 82, 83; Akula, Nirmala 84; McMahon, Francis, J. 84, 85 Binder, Elisabeth, B. 55; Fehm, Lydia 86; Ströhle, Andreas 25; Castelao, Enrique 87; Tiemeier, Henning 88, 89; Stein, Dan, J. 90; Whiteman, David 91; Olsen, Catherine 91; Fuller, Zachary 92; Wang, Xin 92; Wray, Naomi, R. 61, 93; Byrne, Enda, M. 80; Lewis, Glyn 94; Timpson, Nicholas, J. 52, 14; Davis, Lea, K. 17; Hickie, Ian, B. 95; Gillespie, Nathan, A. 6; Milani, Lili 20; Schumacher, Johannes 24; Woldbye, David, P. 96; Ströhle, Andreas 97; Forstner, Andreas, J. 21, 98, 24; Nöthen, Markus, M. 21; Hovatta, Iiris 99; Horwood, John 79; Copeland, William, E. 100; Maes, Hermine, H. 11, 6, 101; McIntosh, Andrew, M. 30; Andreassen, Ole, A. 41, 42, 102; Zwart, John-Anker 44, 37, 45; Mors, Ole 103, 104; Børglum, Anders, D. 4, 104, 105; Mortensen, Preben, B. 106; Ask, Helga 49, 7; Reichborn-Kjennerud, Ted 49, 41; Najman, Jackob, M. 107; Stein, Murray, B. 108, 109; Gelernter, Joel 53, 110, 111; Milaneschi, Yuri 112; Penninx, Brenda, W. 112; Boomsma, Dorret, I. 113, 114; Maron, Eduard 115, 116; Erhardt-Lehmann, Angelika 55, 117; Rück, Christian 3; Kircher, Tilo, T. 118; Melzig, Christiane, A. 119, 120; Alpers, Georg, W. 121; Arolt, Volker 122; Domschke, Katharina 123, 124; Smoller, Jordan, W. 58, 57; Preisig, Martin 59; Martin, Nicholas, G. 12; Lupton, Michelle, K. 12, 13, 125; Luik, Annemarie, I. 126; Reif, Andreas 127; Grabe, Hans, J. 62; Larsson, Henrik 65, 64; Magnusson, Patrik, K. 64; Oldehinkel, Albertine, J. 128; Hartman, Catharina, A. 128; Breen, Gerome 8; Docherty, Anna, R. 129, 130, 6; Coon, Hilary 129; Conrad, Rupert 131; Lehto, Kelli 20; Deckert, Jürgen 117; Eley, Thalia, C. 8; Mattheisen, Manuel 132, 133, 2; Hettema, John, M. 134 1: Department of Psychology, Humboldt-Universität zu Berlin, Berlin, Germany; 2: Institute of Psychiatric Phenomics and Genomics (IPPG), University Hospital, LMU Munich, Munich, Germany; 3: Centre for Psychiatry Research, Department of Clinical Neuroscience, Karolinska Institutet & Stockholm Health Care Services, Region Stockholm, Stockholm, Sweden; 4: Department of Biomedicine, Aarhus University, Aarhus, Denmark; 5: Psychiatry and Behavioral Sciences, Texas A&M University, College Station, Texas, USA; 6: Psychiatry, Virginia Commonwealth University, Richmond, Virginia, USA; 7: PROMENTA Centre, Department of Psychology, University of Oslo, Oslo, Norway; 8: Social, Genetic and Developmental Psychiatry Centre, Institute of Psychiatry, Psychology and Neuroscience, King’s College London, London, UK; 9: Institute for Genomics in Health, Department of Psychiatry and Behavioral Sciences, State University of New York Downstate Health Sciences University, Brooklyn, New York, USA; 10: Life Sciences, Integrative Life Sciences Doctoral Program, Virginia Commonwealth University, Richmond, Virginia, USA; 11: Human and Molecular Genetics, Virginia Institute for Psychiatric and Behavioral Genetics, Virginia Commonwealth University, Richmond, Virginia, USA; 12: Brain and Mental Health Program, QIMR Berghofer Medical Research Institute, Brisbane, Queensland, Australia; 13: Faculty of Medicine, Queensland University, Brisbane, Queensland, Australia; 14: Bristol Medical School, Population Health Sciences, MRC Integrative Epidemiology Unit, University of Bristol, Bristol, UK; 15: Centre for Clinical Brain Sciences, Division of Psychiatry, University of Edinburgh, Edinburgh, UK; 16: Division of Medicine, Human Genetics, Vanderbilt University, Nashville, Tennessee, USA; 17: Division of Genetic Medicine, Vanderbilt University Medical Center, Nashville, Tennessee, USA; 18: Vanderbilt Genetics Institute, Vanderbilt University Medical Center, Nashville, Tennessee, USA; 19: School of Psychology, The University of Queensland, Brisbane, Queensland, Australia; 20: Estonian Genome Centre, Institute of Genomics, University of Tartu, Tartu, Estonia; 21: Institute of Human Genetics, University of Bonn, School of Medicine & University Hospital Bonn, Bonn, Germany; 22: Department of Biomedicine, Human Genomics Research Group, University of Basel; University Hospital Basel, Basel, Switzerland; 23: Institute of Medical Genetics and Pathology, Medical Faculty, University Hospital Basel, Basel, Switzerland; 24: Center for Human Genetics, University of Marburg, Marburg, Germany; 25: Dept. of Psychiatry and Psychotherapy, Charité - Universitätsmedizin, Berlin, Germany; 26: Analytic and Translational Genetics Unit, Massachusetts General Hospital, Boston, Massachusetts, USA; 27: Helsinki Institute of Life Science, Institute for Molecular Medicine Finland - FIMM, University of Helsinki, Helsinki, Finland; 28: Faculty of Medicine, Department of Psychology and Logopedics, SleepWell Research Program, University of Helsinki, Helsinki, Finland; 29: School of Medicine, Department of Psychiatry, University of Utah, Salt Lake City, Utah, USA; 30: Centre for Clinical Brain Sciences, University of Edinburgh, Edinburgh, UK; 31: College of Medicine and Veterinary Medicine, Institute of Genetics and Cancer; Centre for Genomic and Experimental Medicine, University of Edinburgh, Edinburgh, UK; 32: Department of Clinical and Molecular Medicine, Norwegian University of Science and Technology, Trondheim, Norway; 33: HUNT Center for Molecular and Clinical Epidemiology, Department of Public Health and Nursing, Faculty of Medicine and Health Sciences, Norwegian University of Science and Technology, Trondheim, Norway; 34: BioCore - Bioinformatics Core Facility, Norwegian University of Science and Technology, Trondheim, Norway; 35: Clinic of Laboratory Medicine, St. Olavs Hospital, Trondheim University Hospital, Trondheim, Norway; 36: Division of Clinical Neuroscience, Department of Research and Innovation, Oslo University Hospital, Oslo, Norway; 37: Department of Public Health and Nursing, HUNT Center for Molecular and Clinical Epidemiology, Norwegian University of Science and Technology, Trondheim, Norway; 38: Department of Neurology, Oslo University Hospital, Oslo, Norway; 39: Department of Mental Health, Norwegian University of Science and Technology, Trondheim, Norway; 40: Division of Mental Health, St. Olavs Hospital, Trondheim University Hospital, Trondheim, Norway; 41: NORMENT Centre, University of Oslo, Oslo, Norway; 42: Centre of Precision Psychiatry, Division of Mental Health and Addiction, Oslo University Hospital and University of Oslo, Oslo, Norway; 43: Department of Psychiatry, Sørlandet Hospital, Kristiansand, Norway; 44: Division of Clinical Neuroscience, Department of Research and Innovation; Musculoskeletal Health, Oslo University Hospital, Oslo, Norway; 45: Faculty of Medicine, Institute of Clinical Medicine, University of Oslo, Oslo, Norway; 46: National Institute for Health and Care Research (NIHR) Maudsley Biomedical Research Centre, South London and Maudsley NHS Foundation Trust, London, UK; 47: Department of Clinical, Educational and Health Psychology, University College London, London, United Kingdom; 48: Psychiatry, Dalhousie University, Halifax, Nova Scotia, Canada; 49: PsychGen Centre for Genetic Epidemiology and Mental Health, Norwegian Institute of Public Health, Oslo, Norway; 50: Nic Waals Institute, Lovisenberg Diaconal Hospital, Oslo, Norway; 51: Nic Waals Institute, Lovisenberg Diaconal Hospital, Oslo, Norway; 52: Bristol Medical School, Population Health Sciences, University of Bristol, Bristol, UK; 53: Department of Psychiatry, Division of Human Genetics, Yale University School of Medicine, New Haven, Connecticut, USA; 54: Psychiatry, Research, Veterans Affairs Connecticut Healthcare System, West Haven, Connecticut, USA; 55: Department of Genes and Environment, Max-Planck Institute of Psychiatry, Munich, Germany; 56: Department of Psychiatry, Psychosomatics and Psychotherapy, University Hospital of Würzburg, Würzburg, Germany; 57: Psychiatry, Center for Precision Psychiatry, Massachusetts General Hospital, Boston, Massachusetts, USA; 58: Psychiatry, Psychiatric and Neurodevelopmental Genetics Unit, Center for Genomic Medicine, Massachusetts General Hospital, Boston, Massachusetts, USA; 59: Psychiatric Epidemiology and Psychopathology Research Center, Department of Psychiatry, Lausanne University Hospital and University of Lausanne, Prilly, Switzerland; 60: ARAMIS laboratory, Paris Brain Institute, Paris, France; 61: Institute for Molecular Bioscience, University of Queensland, Brisbane, Queensland, Australia; 62: Department of Psychiatry and Psychotherapy, University Medicine Greifswald, Greifswald, Germany; 63: Institute for Community Medicine, University Medicine Greifswald, Greifswald, Germany; 64: Department of Medical Epidemiology and Biostatistics, Karolinska Institutet, Stockholm, Sweden; 65: School of Medical Sciences, Faculty of Medicine and Health, Örebro University, Örebro, Sweden; 66: Department of Statistics, Miami University, Oxford, Ohio, USA; 67: Social, Genetic, and Developmental Psychiatry Centre, Institute of Psychiatry, Psychology and Neuroscience, King’s College London, London, UK; 68: Faculty of Nursing and Health Science, Nord University, Levanger, Norway; 69: Department of Psychiatry, Hospital Namsos, Nord-Trøndelag Health Trustt, Namsos, Norway; 70: Department of Psychiatry, Universidade Federal do Rio Grande do Sul, Porto Alegre, Rio Grande do Sul, Brazil; 71: Child Psychiatry, National Institute of Developmental Psychiatry, São Paulo, Brazil; 72: Tanenbaum Centre for Pharmacogenetics, Molecular Brain Sciences Department, Campbell Family Mental Health Institute, Centre for Addiction and Mental Health, Toronto, Ontario, Canada; 73: Department of Psychiatry, Division of Neurosciences and Clinical Translation, University of Toronto, Toronto, Ontario, Canada; 74: Institute of Medical Science, University of Toronto, Toronto, Ontario, Canada; 75: Laboratory Medicine and Pathobiology, University of Toronto, Toronto, Ontario, Canada; 76: Stanley Center for Psychiatric Research, Broad Institute of Harvard and MIT, Cambridge, MA, USA; 77: Institute for Medical Informatics, Biometry and Epidemiology, University Hospital of Essen, University of Duisburg-Essen, Essen, Germany; 78: Pathology and Biomedical Science, University of Otago, Christchurch, New Zealand; 79: Psychological Medicine, University of Otago, Christchurch, New Zealand; 80: Child Health Research Centre, University of Queensland, Brisbane, Queensland, Australia; 81: Child and Youth Mental Health Service, Children’s Health Queensland Hospital and Health Service, Brisbane, Queensland, Australia; 82: National Institute of Mental Health, Human Genetics Branch, National Institutes of Health, Bethesda, Maryland, USA; 83: Department of Psychiatry and Human Behavior, Alpert Medical School of Brown University, Providence, Rhode Island, USA; 84: National Institute of Mental Health, Genetic Basis of Mood and Anxiety Disorders, National Institutes of Health, Bethesda, Maryland, USA; 85: Psychiatry & Behavioral Sciences, Johns Hopkins University, Baltimore, Maryland, USA; 86: Department of Psychology, Zentrum für Psychotherapie, Humboldt-Universität zu Berlin, Berlin, Germany; 87: Psychiatric Epidemiology and Psychopathology Research Center, Department of Psychiatry, Lausanne University Hospital and University of Lausanne, Prilly, Switzerland; 88: Social and Behavioral Science, T.H. Chan School of Public Health, Harvard University, Boston, Massachusetts, USA; 89: Child and Adolescent Psychiatry, Erasmus University Medical Center, Rotterdam, Netherlands; 90: SAMRC Unit on Risk & Resilience in Mental Disorders, Department of Psychiatry & Neuroscience Institute, University of Cape Town, Cape Town, South Africa; 91: Population Health Program, QIMR Berghofer Medical Research Institute, Brisbane, Australia; 92: 23andMe, Sunnyvale, CA, USA; 93: Department of Psychiatry, University of Oxford, Oxford, UK; 94: UCL Division of Psychiatry, University College London, London, UK; 95: Brain and Mind Centre, University of Sydney, Sydney, Australia; 96: Department of Neuroscience, Laboratory of Neural Plasticity, University of Copenhagen, Copenhagen, Denmark; 97: Department of Psychiatry and Psychotherapy, Campus Charité Mitte, Charité - Universitätsmedizin Berlin, Corporate member of Freie Universität Berlin and Humboldt-Universität zu Berlin, Berlin, Germany; 98: Institute of Neuroscience and Medicine (INM-1), Research Center Jülich, Jülich, Germany; 99: Faculty of Medicine, Department of Psychology and Logopedics and SleepWell Research Program, University of Helsinki, Helsinki, Finland; 100: UVM Medical Center, Department of Psychiatry, University of Vermont, Burlington, Vermont, USA; 101: Massey Cancer Center, Virginia Commonwealth University, Richmond, Virginia, USA; 102: K. G. Jebsen Center for Neurodevelopmental disorders, University of Oslo, Oslo, Norway; 103: Department of Psychiatry, Psychosis Research Unit, Aarhus University Hospital, Aarhus, Denmark; 104: The Lundbeck Foundation Initiative for Integrative Psychiatric Research, iPSYCH, Aarhus University, Aarhus, Denmark; 105: Center for Genomics and Personalised Medicine, Aarhus University, Aarhus, Denmark; 106: The National Centre for Register-based Research, Aarhus University, Aarhus, Denmark; 107: Faculty of Medicine, School of Public Health, University of Queensland, Herston, Queensland, Australia; 108: Psychiatry, University of California San Diego, La Jolla, CA, USA; 109: School of Public Health, University of California San Diego, La Jolla, CA, USA; 110: Psychiatry Research, Veterans Affairs Connecticut Healthcare System, West Haven, Connecticut, USA; 111: Departments of Genetics and Neuroscience, Yale University of Medicine, New Haven, Connecticut, USA; 112: Amsterdam Neuroscience; Amsterdam Public Health, Amsterdam University Medical Center, Amsterdam, Netherlands; 113: Twin Register and Department of Complex Trait Genetics, Center for Neurogenomics and Cognitive Research, Vrije Universiteit Amsterdam, Amsterdam, Netherlands; 114: Amsterdam Public Health, Amsterdam University Medical Center, Amsterdam, Netherlands; 115: Psychiatry, University of Tartu, Tartu, Estonia; 116: Department of Medicine, Centre for Neuropsychopharmacology,, Division of Brain Sciences, Imperial College London, London, UK; 117: Department of Psychiatry, Psychosomatics and Psychotherapy, University Hospital Würzburg, Würzburg, Germany; 118: Department of Psychiatry, University of Marburg, Marburg, Germany; 119: Psychology, Clinical Psychology, Experimental Psychopathology and Psychotherapy, University of Marburg, Marburg, Germany; 120: Psychology, Biological and Clinical Psychology, University of Greifswald, Greifswald, Germany; 121: School of Social Sciences, Department of Psychology, University of Mannheim, Mannheim, Germany; 122: Department of Mental Health, Institute for Translational Psychiatry, University of Muenster, Muenster, Germany; 123: Department of Psychiatry and Psychotherapy, Medical Center, Faculty of Medicine, University of Freiburg, Freiburg, Germany; 124: German Center for Mental Health (DZPG), Parnter Site Berlin, Berlin, Germany; 125: Faculty of Health, Queensland University of technology, Queensland, Australia; 126: Epidemiology, Erasmus University Medical Center, Rotterdam, Netherlands; 127: Department of Psychiatry, Psychosomatic Medicine and Psychotherapy, University Hospital Frankfurt - Goethe University, Frankfurt, Germany; 128: Psychiatry, Interdisciplinary Center Psychopathology and Emotion Regulation, University of Groningen, University Medical Center Groningen, Groningen, Netherlands; 129: School of Medicine, Psychiatry, University of Utah, Salt Lake City, Utah, USA; 130: School of Medicine, Psychiatry; Huntsman Mental Health Institute, University of Utah, Salt Lake City, Utah, USA; 131: Department of Psychosomatic Medicine and Psychotherapy, University Hospital Münster, Münster, Germany; 132: Community Health and Epidemiology, Dalhousie University, Halifax, Nova Scotia, Canada; 133: Computer Science, Dalhousie University, Halifax, Nova Scotia, Canada; 134: Psychiatry and Behavioral Sciences, Texas A&M University, Bryan, Texas, USA

**GLAD+ group author:** Gerome Breen, Thalia C. Eley, Matthew Hotopf, Chérie Armour, Ian R. Jones, Andrew M. McIntosh, James T. R. Walters, Daniel J. Smith, Jonathan R. I. Coleman, Allan H. Young, Antony J. Cleare, Katrina A. S. Davis, Georgina Krebs, Katharine A. Rimes, David Veale, Roland Zahn, Molly R. Davies, Gursharan Kalsi, Shannon Bristow, Susannah C. B. Curzons, Henry C. Rogers, Katherine N. Thompson, Brett N. Adey, Ian Marsh, Dina Monssen, Monika McAtarsney-Kovacs, Charles J. Curtis, Jahnavi Arora, Saakshi Kakar, Laura Meldrum, Iona Smith, Le Roy Dowey, Victor Gault, Donald M. Lyall, Ruth K. Price, Keith G. Thomas, Zain Ahmad, Helena L. Davies, Christopher Hübel, Sang Hyuck Lee, Abigail R. ter Kuile, Danyang Li, Yuhao (Leo) Lin, Jared G. Maina, Jessica Mundy, Alish Palmos, Alicia J. Peel, Kirstin Purves, Christopher Rayner, Megan Skelton, Rujia Wang, Johan Zvrskovec

**UMCG Genetics Lifelines Initiative (UGLI) group author:** LifeLines Cohort Study - Raul Aguirre-Gamboa, Patrick Deelen, Lude Franke, Jan A Kuivenhoven, Esteban A Lopera Maya, Ilja M Nolte, Serena Sanna, Harold Snieder, Morris A Swertz, Peter M. Visscher, Judith M Vonk, Cisca Wijmenga, Naomi Wray. See supplemental information for consortium member affiliations.

**NIHR BioResource group author:** John Bradley, Nathalie Kingston, Hannah Stark, Carola Kanz Alexei Moulton, Nigel Ovington, Jacinta Lee, Debbie Clapham-Riley, Katie Mills

**PROTECT-AD:** Principal investigators and responsible for recruitment are Uli Wittchen, Andre Pittig, Ingmar Heinig, Jürgen Hoyer (Dresden), Andreas Ströhle and Jens Fydrich (Berlin), Alfons Hamm and Jan Richter (Greifswald), Volker Arolt, Udo Dannlowski and Katja Kölkebeck (Münster), Sylvia Schneider and Jürgen Margraf (Bochum), Tilo Kircher, Benjamin Straube and Wilfried Rief (Marburg), Jürgen Deckert, Katharina Domschke, Ulrike Lueken and Paul Pauli (Würzburg) and Peter Neudeck (Cologne). The presented work was derived from projects P1 and P5. We would like to thank the following individuals for their help: Jule Dehler, Dorte Westphal, Katrin Hummel, Jürgen Hoyer (Dresden), Verena Pflug, Dirk Adolph, Cornelia Mohr, Jan Cwik (Bochum), Maike Hollandt, Anne Pietzner, Jörg Neubert (Greifswald), Carsten Konrad, Yunbo Yang, Isabelle Ridderbusch, Adrian Wroblewski, Hanna Christiansen, Anne Maenz, Sophia Tennie, Jean Thierschmidt (Marburg), Marcel Romanos, Kathrin Zierhut, Kristina Dickhöver, Markus Winkler, Maria Stefanescu, Christiane Ziegler, Heike Weber (Würzburg), Nathalia Weber, Sebastian Schauenberg, Sophia Wriedt, Carina Heitmann (Münster) Caroline im Brahm, Annika Evers (Cologne), Isabel Alt, Sophie Bischoff, Jennifer Mumm, Jens Plag, Anne Schreiner, Sophie Meska (Berlin). Xina Grählert and Marko Käppler of the Coordinating Centre for Clinical Trials (KKS) data center (Dresden) provided support with the electronic data assessment and data banking. Eva Stolzenburg, Stanislav Bologov, and Karina Bley provided administrative support.

## Notes

### Author Declarations

The study received ethical approval by the QIMR Berghofer Medical Research Institute Human Research Ethics Committee in Brisbane, Australia. Written informed consent was obtained from participants. Ethical approval was obtained from the ALSPAC Ethics and Law Committee and the Local Research Ethics Committees. The ALSPAC biorepository is an NHS Research Ethics Committee (REC) approved Research Tissue Bank (RTB), REC reference 23/SW/0058. This research was approved by the local institutional review board. All participants received a detailed description of the goal and funding of the study and signed a written consent. The activities of the Estonian Biobank are regulated by the Human Genes Research Act, which was adopted in 2000 specifically for the operations of the Estonian Biobank. In this study, analysis of individual level data of the Estonian Biobank was carried out under ethical approval nr 1.1-12/624 and its extensions from the Estonian Committee on Bioethics and Human Research (Estonian Ministry of Social Affairs). All actions are in concert with the General Data Protection Regulation of the European Union. Ethical approval for the original data collection was obtained from the Tayside Committee on Medical Research Ethics (ref 05/S1401/89). Generation Scotland is currently approved as a Research Tissue Bank by the East of Scotland Research Ethics Service (ref 20/ES/0021). Ethical approval for the GLAD Study and NBR COPING study was obtained from the London-Fulham Research Ethics Committee (REC reference 18/LO/1218 and 20/SW/0078, respectively, COPING project no. 282754.). The dataset used in the present analysis was freeze 2023-06-07. All participants provided informed consent. The Lifelines Cohort study is conducted according to the principles of the Declaration of Helsinki and in accordance with the research code of University Medical Center Groningen, and is approved by its medical ethical committee. All participants signed an informed consent form. CRC TRR-58 was approved by the ethics committee of the University of Wurzburg (project no. 304/15). All participants provided informed consent, and the study protocol was approved by the Veterans Affairs Central Institutional Review Board. Summary statistics obtained through dbGaP (phs001672). The establishment of MoBa and initial data collection was based on a license from the Norwegian Data Protection Agency and approval from The Regional Committees for Medical and Health Research Ethics. The MoBa cohort is now based on regulations related to the Norwegian Health Registry Act. The current study was approved by The Regional Committees for Medical and Health Research Ethics (20311). The Norwegian Mother, Father and Child Cohort Study is supported by the Norwegian Ministry of Health and Care Services and the Ministry of Education and Research. The study was performed according to the Declaration of Helsinki and was approved by the Ethics Committee of Technische Universitat Dresden, Germany (EK 234062014). TEDS has been granted ethical approval by the Kings College London ethics committee (References: PNM/09/10-104 and HR/DP-20/21-22060). Informed consent was obtained from all participants. The study was approved by the Dutch Central Committee on Research Involving Human Subjects. Participants were treated in compliance with the Declaration of Helsinki, and all measurements were carried out with their adequate understanding and written consent. UKB analyses were conducted under application ID 82087. All participants provided informed consent.

### Summary of Updates

This manuscript has been rivised which has resulted in slight changes to the overall sample size and therefore the point estimates of downstream analyses

## References

1. GBD 2021 Diseases and Injuries Collaborators. Global incidence, prevalence, years lived with disability (YLDs), disability-adjusted life-years (DALYs), and healthy life expectancy (HALE) for 371 diseases and injuries in 204 countries and territories and 811 subnational locations, 1990-2021: a systematic analysis for the Global Burden of Disease Study 2021. Lancet 403, 2133–2161 (2024).

2. Archer, C., Turner, K., Kessler, D., Mars, B. & Wiles, N. Trends in the recording of anxiety in UK primary care: a multi-method approach. Soc. Psychiatry Psychiatr. Epidemiol. 57, 375–386 (2022).

3. Goodwin, R. D., Weinberger, A. H., Kim, J. H., Wu, M. & Galea, S. Trends in anxiety among adults in the United States, 2008-2018: Rapid increases among young adults. J. Psychiatr. Res. 130, 441–446 (2020).

4. GBD 2019 Mental Disorders Collaborators. Global, regional, and national burden of 12 mental disorders in 204 countries and territories, 1990-2019: a systematic analysis for the Global Burden of Disease Study 2019. Lancet Psychiatry 9, 137–150 (2022).

5. Rapaport, M. H., Clary, C., Fayyad, R. & Endicott, J. Quality-of-Life Impairment in Depressive and Anxiety Disorders. American Journal of Psychiatry 162, 1171–1178 (2005).

6. Meier, S. M. & Deckert, J. Genetics of Anxiety Disorders. Curr. Psychiatry Rep. 21, 16 (2019).

7. Pratt, L. A., Druss, B. G., Manderscheid, R. W. & Walker, E. R. Excess mortality due to depression and anxiety in the United States: Results from a nationally representative survey. Gen. Hosp. Psychiatry 39, 39–45 (2016).

8. Wittchen, H. U., Zhao, S., Kessler, R. C. & Eaton, W. W. DSM-III-R generalized anxiety disorder in the National Comorbidity Survey. Arch. Gen. Psychiatry 51, 355–364 (1994).

9. Pavlova, B., Perlis, R. H., Alda, M. & Uher, R. Lifetime prevalence of anxiety disorders in people with bipolar disorder: a systematic review and meta-analysis. Lancet Psychiatry 2, 710–717 (2015).

10. Kessler, R. C., Chiu, W. T., Demler, O., Merikangas, K. R. & Walters, E. E. Prevalence, severity, and comorbidity of 12-month DSM-IV disorders in the National Comorbidity Survey Replication. Arch. Gen. Psychiatry 62, 617–627 (2005).

11. Hollocks, M. J., Lerh, J. W., Magiati, I., Meiser-Stedman, R. & Brugha, T. S. Anxiety and depression in adults with autism spectrum disorder: a systematic review and meta-analysis. Psychol. Med. 49, 559–572 (2019).

12. Achim, A. M. et al. How prevalent are anxiety disorders in schizophrenia? A meta-analysis and critical review on a significant association. Schizophr. Bull. 37, 811–821 (2011).

13. Ask, H. et al. Genetic contributions to anxiety disorders: where we are and where we are heading. Psychol. Med. 51, 2231–2246 (2021).

14. Ozsivadjian, A., Knott, F. & Magiati, I. Parent and child perspectives on the nature of anxiety in children and young people with autism spectrum disorders: a focus group study. Autism 16, 107– 121 (2012).

15. Chadwick, P., Kaur, H., Swelam, M., Ross, S. & Ellett, L. Experience of mindfulness in people with bipolar disorder: a qualitative study. Psychother. Res. 21, 277–285 (2011).

16. Cheesman, R. et al. Extracting stability increases the SNP heritability of emotional problems in young people. Transl. Psychiatry 8, 223 (2018).

17. Otowa, T. et al. Meta-analysis of genome-wide association studies of anxiety disorders. Mol. Psychiatry 21, 1391–1399 (2016).

18. Meier, S. M. et al. Genetic Variants Associated With Anxiety and Stress-Related Disorders: A Genome-Wide Association Study and Mouse-Model Study. JAMA Psychiatry 76, 924–932 (2019).

19. Purves, K. L. et al. A Major Role for Common Genetic Variation in Anxiety Disorders. Mol. Psychiatry 25, 3292–3303 (2020).

20. Li, W. et al. Genome-wide meta-analysis, functional genomics and integrative analyses implicate new risk genes and therapeutic targets for anxiety disorders. Nat. Hum. Behav. 8, 361–379 (2024).

21. Verma, A. et al. Diversity and scale: Genetic architecture of 2068 traits in the VA Million Veteran Program. Science 385, eadj1182 (2024).

22. Friligkou, E. et al. Gene discovery and biological insights into anxiety disorders from a large-scale multi-ancestry genome-wide association study. Nat. Genet. 1–10 (2024).

23. Strom, N. I. et al. Genome-wide association study of major anxiety disorders in 122,341 European-ancestry cases identifies 58 loci and highlights GABAergic signaling. bioRxiv (2024) doi:10.1101/2024.07.03.24309466.

24. Lee, W. E., Wadsworth, M. E. J. & Hotopf, M. The protective role of trait anxiety: a longitudinal cohort study. Psychol. Med. 36, 345–351 (2006).

25. Altman, D. G. & Royston, P. The cost of dichotomising continuous variables. BMJ 332, 1080 (2006).

26. Levey, D. F. et al. Reproducible Genetic Risk Loci for Anxiety: Results From ⍰200,000 Participants in the Million Veteran Program. Am. J. Psychiatry 177, 223–232 (2020).

27. Skelton, M. et al. Genetic overlap between functional impairment and depression and anxiety symptom severity: Evidence from the GLAD Study. PsyArXiv (2025) doi:10.31234/osf.io/4d6ju_v2.

28. Tesfaye, M. et al. Identification of novel genomic loci for anxiety symptoms and extensive genetic overlap with psychiatric disorders. Psychiatry Clin. Neurosci. 78, 783–791 (2024).

29. Thorp, J. G. et al. Symptom-level modelling unravels the shared genetic architecture of anxiety and depression. Nature Human Behaviour 5, 1432–1442 (2021).

30. Visscher, P. M. et al. Statistical power to detect genetic (co)variance of complex traits using SNP data in unrelated samples. PLoS Genet. 10, e1004269 (2014).

31. Evans, L. M. & Keller, M. C. Using partitioned heritability methods to explore genetic architecture. Nat. Rev. Genet., 185 (2018).

32. Bulik-Sullivan, B. et al. LD Score regression distinguishes confounding from polygenicity in genome-wide association studies. Nat. Genet. 47, 291–295 (2015).

33. Bulik-Sullivan, B. et al. An atlas of genetic correlations across human diseases and traits. Nat. Genet. 47, 1236–1241 (2015).

34. Zhu, Z. et al. Causal associations between risk factors and common diseases inferred from GWAS summary data. Nat. Commun. 9, 224 (2018).

35. Zheng, Z. et al. Leveraging functional genomic annotations and genome coverage to improve polygenic prediction of complex traits within and between ancestries. Nat. Genet. 56, 767–777 (2024).

36. Weissbrod, O. et al. Functionally informed fine-mapping and polygenic localization of complex trait heritability. Nat. Genet. 52, 1355–1363 (2020).

37. Wang, G., Sarkar, A., Carbonetto, P. & Stephens, M. A simple new approach to variable selection in regression, with application to genetic fine mapping. J. R. Stat. Soc. Series B Stat. Methodol. 82, 1273–1300 (2020).

38. Amendola, L. M. et al. Actionable exomic incidental findings in 6503 participants: challenges of variant classification. Genome Res. 25, 305–315 (2015).

39. Watanabe, K., Taskesen, E., van Bochoven, A. & Posthuma, D. Functional mapping and annotation of genetic associations with FUMA. Nat. Commun. 8, 1826 (2017).

40. Li, Y. et al. Cross-ancestry genome-wide association study and systems-level integrative analyses implicate new risk genes and therapeutic targets for depression. Nat. Hum. Behav. 9, 806–823 (2025).

41. de Leeuw, C. A., Mooij, J. M., Heskes, T. & Posthuma, D. MAGMA: generalized gene-set analysis of GWAS data. PLoS Comput. Biol. 11, e1004219 (2015).

42. Subramanian, A. et al. Gene set enrichment analysis: a knowledge-based approach for interpreting genome-wide expression profiles. Proc. Natl. Acad. Sci. U. S. A. 102, 15545–15550 (2005).

43. GTEx Consortium. The GTEx Consortium atlas of genetic regulatory effects across human tissues. Science 369, 1318–1330 (2020).

44. Gaspar, H. A., Hübel, C. & Breen, G. Drug Targetor: a web interface to investigate the human druggome for over 500 phenotypes. Bioinformatics 35, 2515–2517 (2019).

45. Ren, L. et al. Anxiety disorders: Treatments, models, and circuitry mechanisms. Eur. J. Pharmacol. 983, 176994 (2024).

46. Carey, C. E. et al. Principled distillation of UK Biobank phenotype data reveals underlying structure in human variation. Nat. Hum. Behav. 8, 1599–1615 (2024).

47. Sullivan, P. F. et al. Genome-wide association for major depressive disorder: a possible role for the presynaptic protein piccolo. Mol. Psychiatry 14, 359–375 (2009).

48. Christiansen, G. B. et al. The sorting receptor SorCS3 is a stronger regulator of glutamate receptor functions compared to GABAergic mechanisms in the hippocampus. Hippocampus 27, 235–248 (2017).

49. Breiderhoff, T. et al. Sortilin-related receptor SORCS3 is a postsynaptic modulator of synaptic depression and fear extinction. PLoS One 8, e75006 (2013).

50. Yang, Z. et al. Investigating Shared Genetic Basis Across Tourette Syndrome and Comorbid Neurodevelopmental Disorders Along the Impulsivity-Compulsivity Spectrum. Biol. Psychiatry 90, 317–327 (2021).

51. Sun, F. et al. Hippocampal gray matter volume alterations in patients with first-episode and recurrent major depressive disorder and their associations with gene profiles. BMC Psychiatry 25, 134 (2025).

52. Yang, J., Wray, N. R. & Visscher, P. M. Comparing apples and oranges: equating the power of case-control and quantitative trait association studies. Genet. Epidemiol. 34, 254–257 (2010).

53. Direk, N. et al. An Analysis of Two Genome-wide Association Meta-analyses Identifies a New Locus for Broad Depression Phenotype. Biol. Psychiatry 82, 322–329 (2017).

54. Cai, N. et al. Minimal phenotyping yields genome-wide association signals of low specificity for major depression. Nat. Genet. 52, 437–447 (2020).

55. Huang, L. et al. Polygenic Analyses Show Important Differences Between Major Depressive Disorder Symptoms Measured Using Various Instruments. Biol. Psychiatry (2023) doi:10.1016/j.biopsych.2023.11.021.

56. Strom, N. I. et al. Genome-Wide Association Study of Obsessive-Compulsive Symptoms including 33,943 individuals from the general population. Mol. Psychiatry 29, 2714–2723 (2024).

57. Morneau-Vaillancourt, G. et al. The genetic and environmental hierarchical structure of anxiety and depression in the UK Biobank. Depression and anxiety 37, 512–520 (2020).

58. ter Kuile, A. R. et al. Genome-wide genetic overlap between fear-based disorders and generalised anxiety disorder. (2025).

59. Zugman, A. et al. Mega-analysis methods in ENIGMA: The experience of the generalized anxiety disorder working group. Hum. Brain Mapp. 43, 255–277 (2022).

60. Funk, J., Morneau-Vaillancourt, G., Palaiologou, E. & Eley, T. Heritability of generalised anxiety stability - a longitudinal twin study among young adults. Psychol. Med. (2025).

61. Gillespie, N. A. et al. Do the genetic or environmental determinants of anxiety and depression change with age? A longitudinal study of Australian twins. Twin Res. 7, 39–53 (2004).

62. Rosseel, Y. lavaan: AnRPackage for Structural Equation Modeling. J. Stat. Softw. 48, 1–36 (2012).

63. Mitchell, B. L. et al. The Australian Genetics of Depression Study: New Risk Loci and Dissecting Heterogeneity Between Subtypes. Biol. Psychiatry 92, 227–235 (2022).

64. Boyd, A. et al. Cohort Profile: the ‘children of the 90s’--the index offspring of the Avon Longitudinal Study of Parents and Children. Int. J. Epidemiol. 42, 111–127 (2013).

65. Fraser, A. et al. Cohort Profile: the Avon Longitudinal Study of Parents and Children: ALSPAC mothers cohort. Int. J. Epidemiol. 42, 97–110 (2013).

66. Preisig, M. et al. The PsyCoLaus study: methodology and characteristics of the sample of a population-based survey on psychiatric disorders and their association with genetic and cardiovascular risk factors. BMC Psychiatry 9, 9 (2009).

67. Milani, L. et al. The Estonian Biobank’s journey from biobanking to personalized medicine. Nat. Commun. 16, 3270 (2025).

68. Smith, B. H. et al. Cohort Profile: Generation Scotland: Scottish Family Health Study (GS:SFHS). The study, its participants and their potential for genetic research on health and illness. Int. J. Epidemiol. 42, 689–700 (2013).

69. Davies, M. R. et al. The Genetic Links to Anxiety and Depression (GLAD) Study: Online recruitment into the largest recontactable study of depression and anxiety. Behav. Res. Ther. 123, 103503 (2019).

70. Sijtsma, A. et al. Cohort Profile Update: Lifelines, a three-generation cohort study and biobank. Int. J. Epidemiol. 51, e295–e302 (2022).

71. Deckert, J. et al. GLRB allelic variation associated with agoraphobic cognitions, increased startle response and fear network activation: a potential neurogenetic pathway to panic disorder. Mol. Psychiatry 22, 1431–1439 (2017).

72. Magnus, P. et al. Cohort profile update: The Norwegian Mother and Child Cohort Study (MoBa). Int. J. Epidemiol. 45, 382–388 (2016).

73. Heinig, I. et al. Optimizing exposure-based CBT for anxiety disorders via enhanced extinction: Design and methods of a multicentre randomized clinical trial. Int. J. Methods Psychiatr. Res. 26, (2017).

74. Lockhart, C. et al. Twins Early Development Study (TEDS): A genetically sensitive investigation of mental health outcomes in the mid-twenties. JCPP Adv. (2023) doi:10.1002/jcv2.12154.

75. Oldehinkel, A. J. et al. Cohort profile update: The TRacking adolescents’ individual lives survey (TRAILS). Int. J. Epidemiol. 44, 76–76n (2015).

76. Bycroft, C. et al. The UK Biobank resource with deep phenotyping and genomic data. Nature 562, 203–209 (2018).

77. McCarthy, S. et al. A reference panel of 64,976 haplotypes for genotype imputation. Nat. Genet. 48, 1279–1283 (2016).

78. Willer, C. J., Li, Y. & Abecasis, G. R. METAL: fast and efficient meta-analysis of genomewide association scans. Bioinformatics 26, 2190–2191 (2010).

79. Zheng, J. et al. LD Hub: a centralized database and web interface to perform LD score regression that maximizes the potential of summary level GWAS data for SNP heritability and genetic correlation analysis. Bioinformatics 33, 272–279 (2017).

80. Adams, M. J. et al. Trans-ancestry genome-wide study of depression identifies 697 associations implicating cell types and pharmacotherapies. Cell (2025) doi:10.1016/j.cell.2024.12.002.

81. Cerezo, M. et al. The NHGRI-EBI GWAS Catalog: standards for reusability, sustainability and diversity. Nucleic Acids Res. 53, D998–D1005 (2025).

82. Lin, S.-H., Brown, D. W. & Machiela, M. J. LDtrait: An online tool for identifying published phenotype associations in linkage disequilibrium. Cancer Res. 80, 3443–3446 (2020).

83. Quinlan, A. R. & Hall, I. M. BEDTools: a flexible suite of utilities for comparing genomic features. Bioinformatics 26, 841–842 (2010).

84. Gupta, P. et al. A genome-wide investigation into the underlying genetic architecture of personality traits and overlap with psychopathology. Nat. Hum. Behav. 1–15 (2024).

85. Gilchrist, L. et al. Depression symptom-specific genetic associations in clinically diagnosed and proxy case Alzheimer’s disease. Nat. Ment. Health 3, 212–228 (2025).

86. Lupton, M. K. et al. A prospective cohort study of prodromal Alzheimer’s disease: Prospective Imaging Study of Ageing: Genes, Brain and Behaviour (PISA). NeuroImage Clin. 29, 102527 (2021).

87. Lee, S. H., Goddard, M. E., Wray, N. R. & Visscher, P. M. A better coefficient of determination for genetic profile analysis. Genet. Epidemiol. 36, 214–224 (2012).

88. Ramasamy, A. et al. Genetic variability in the regulation of gene expression in ten regions of the human brain. Nat. Neurosci. 17, 1418–1428 (2014).

89. Schmitt, A. D. et al. A compendium of chromatin contact maps reveals spatially active regions in the human genome. Cell Rep. 17, 2042–2059 (2016).

90. Giusti-Rodriguez, P. M. D. & Sullivan, P. F. Using three-dimensional regulatory chromatin interactions from adult and fetal cortex to interpret genetic results for psychiatric disorders and cognitive traits. bioRxiv 406330 (2018) doi:10.1101/406330.

91. Gaulton, A. et al. ChEMBL: a large-scale bioactivity database for drug discovery. Nucleic Acids Res. 40, D1100–7 (2012).

92. Zdrazil, B. et al. The ChEMBL Database in 2023: a drug discovery platform spanning multiple bioactivity data types and time periods. Nucleic Acids Res. 52, D1180–D1192 (2024).

93. Freshour, S. L. et al. Integration of the Drug-Gene Interaction Database (DGIdb 4.0) with open crowdsource efforts. Nucleic Acids Res. 49, D1144–D1151 (2021).

