## Supplementary Figuress for "Genome-wide analyses of quantitative generalised anxiety symptom severity"

### Supplementary figures for "Genome-wide analyses of quantitative generalised anxiety symptom severity"

**Fig. S1 |** Quantile-quantile plot of dimensional anxiety genome-wide meta-analysis

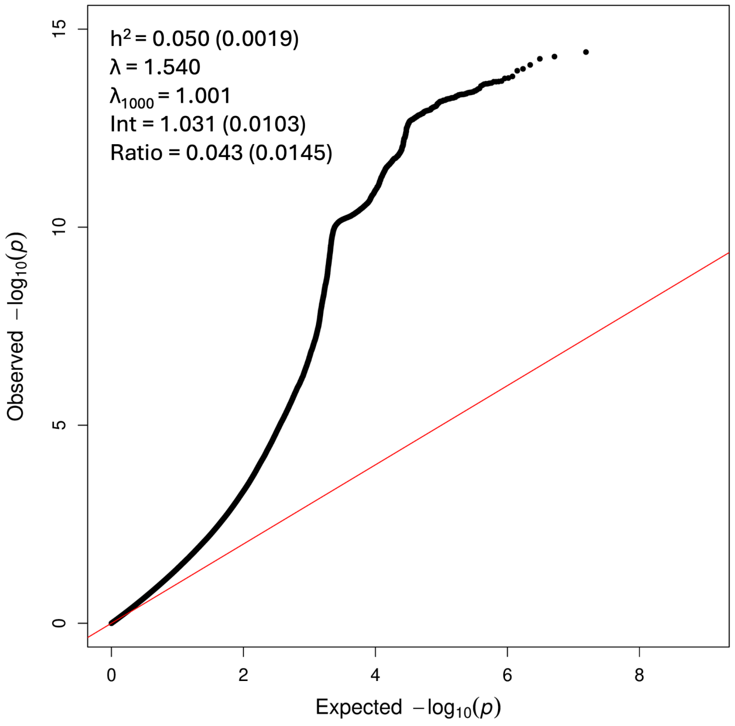


Note: Text is LDSC output, where $h^2$ = heritability, Int = intercept, values in brackets are standard errors of the estimates, $\lambda$ = genomic inflation factor, $\lambda_{1000}$ = genomic inflation factor for an equivalent study with a sample size of 1000.

#### MAGMA Tissue expression results

**Fig. S2 |** BrainSpan 29 different ages of brain samples

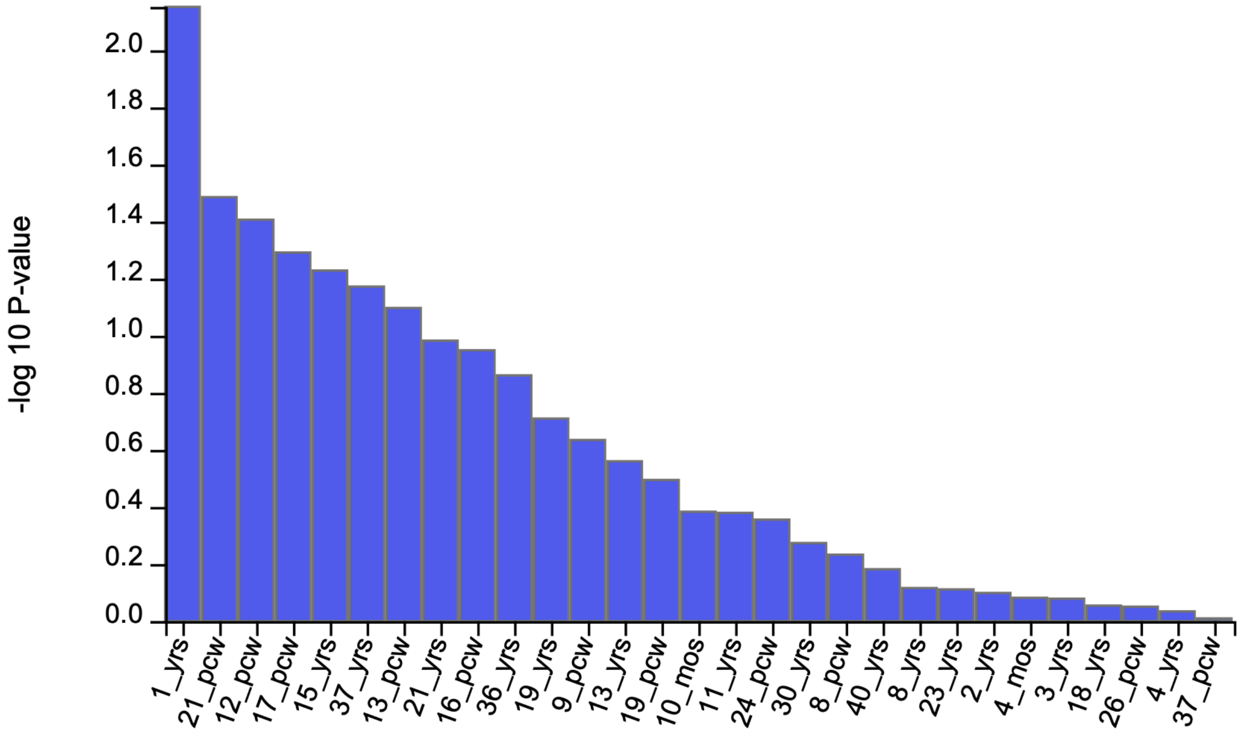


**Fig. S3 |** Gene-tissue results using 11 general developmental stages of brain samples from BrainSpan. The horizontal dotted line indicates the Bonferroni-corrected significance threshold.

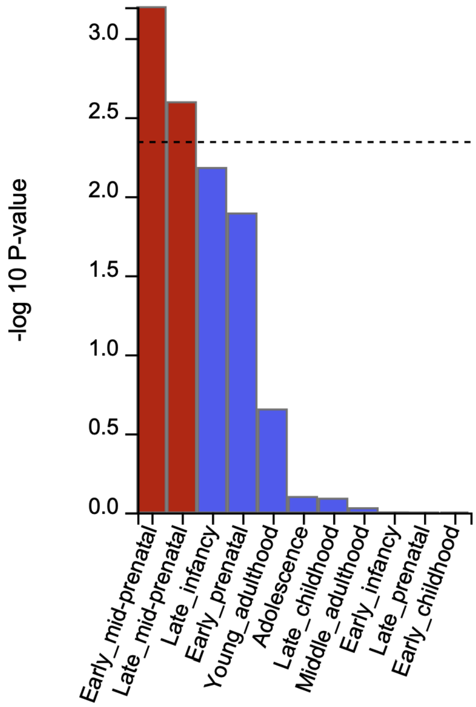

**Fig. S4 |** Gene-tissue results using 30 general tissue types from GTEx v8. The horizontal dotted line indicates the Bonferroni-corrected significance threshold.

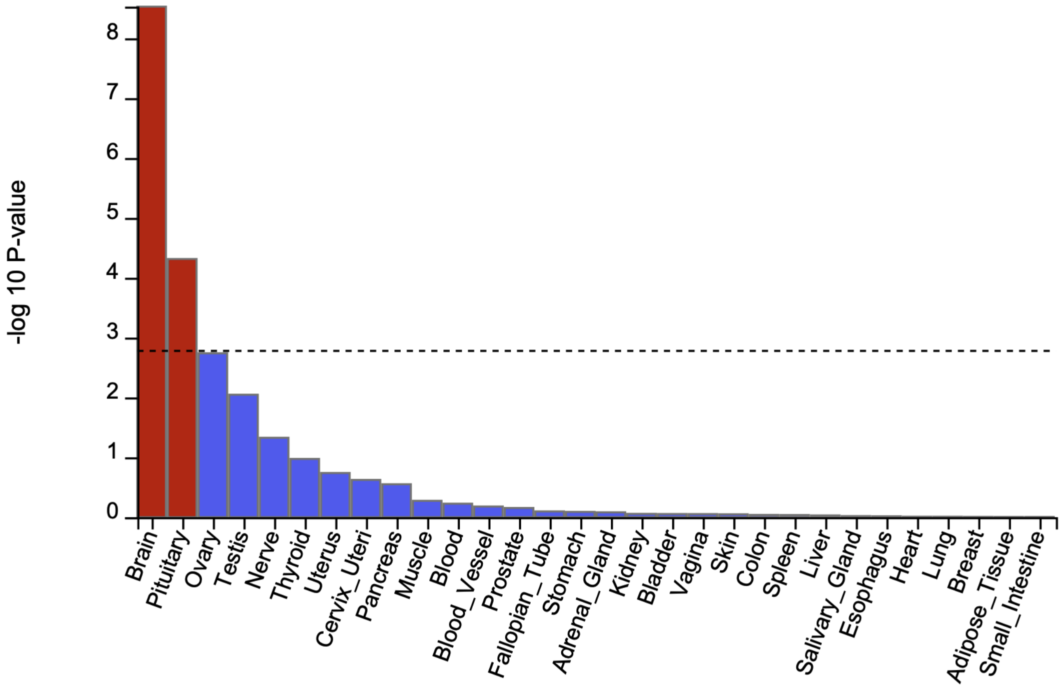


**Fig. S5 |** Gene-tissue results using 53 tissue types from GTEx v8. The horizontal dotted line indicates the Bonferroni-corrected significance threshold.

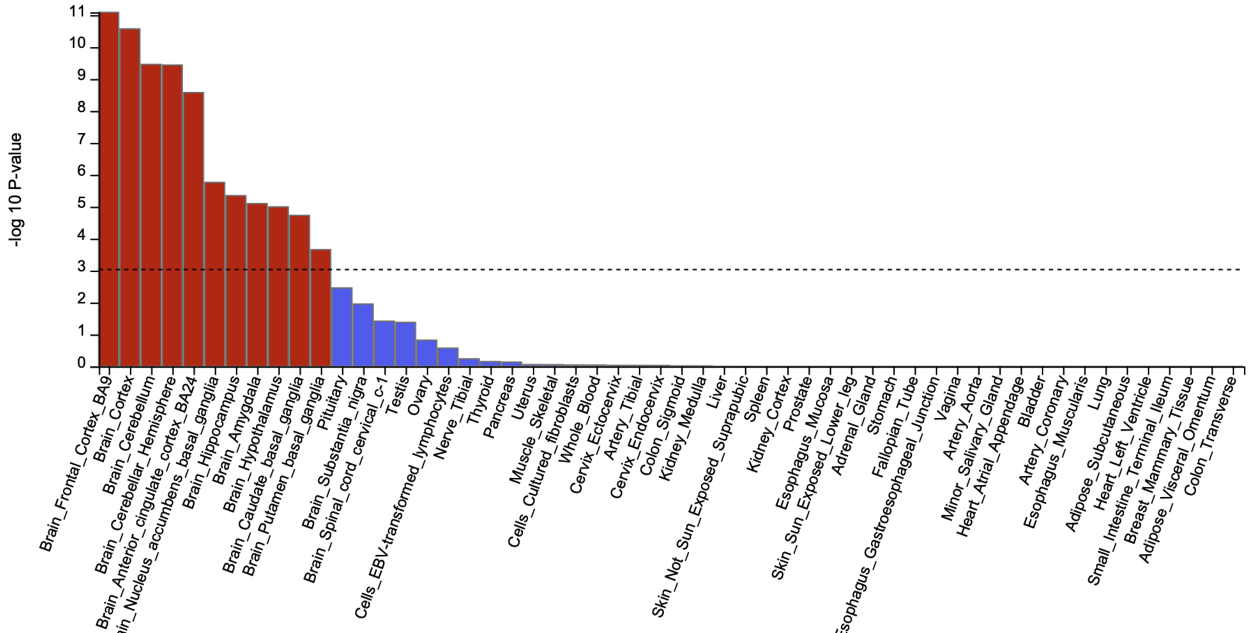
